# *CFTR*-mediated monocyte-macrophage dysfunction revealed by cystic fibrosis proband- parent comparisons

**DOI:** 10.1101/2021.06.30.21259182

**Authors:** Xi Zhang, Camille Moore, Laura Harmacek, Joanne Domenico, Vittobai Rangaraj, Justin E. Ideozu, Jennifer R. Knapp, Kate A. Woods, Stephanie Jump, Shuang Jia, Jeremy W. Prokop, Russel Bowler, Martin J. Hessner, Erwin W. Gelfand, Hara Levy

## Abstract

Cystic fibrosis (CF) is an inherited disorder caused by biallelic mutations of the cystic fibrosis transmembrane conductance regulator gene (*CFTR*). Converging lines of evidence suggest that CF carriers with only one defective *CFTR* copy are at increased risk for CF-related conditions and pulmonary infections, but the molecular mechanisms underpinning this effect remain unknown. Here, we performed transcriptomic profiling of peripheral blood mononuclear cells (PBMCs) of CF child-parent trios (proband, father, and mother) and healthy control PBMCs or THP-1 cells incubated with the plasma of these subjects. Transcriptomic analyses revealed suppression of cytokine-enriched immune-related genes (*IL-1β*, *CXCL8*, *CREM*) implicating lipopolysaccharide tolerance in innate immune cells (monocytes) of CF probands and their parents and in the control innate immune cells incubated with proband or parent plasma. These data suggest that not only a homozygous but also a heterozygous *CFTR* mutation can modulate the immune/inflammatory system. This conclusion is further supported by the findings of lower numbers of circulating monocytes in CF probands and their parents compared to healthy controls, the abundance of mononuclear phagocyte subsets (macrophages, monocytes, and activated dendritic cells) which correlated with *Pseudomonas aeruginosa* infection, lung disease severity, and CF progression in the probands. This study provides insight into demonstrated *CFTR*-related innate immune dysfunction in individuals with CF and carriers of a *CFTR* mutation that may serve as a target for personalized therapy.

## Introduction

Cystic fibrosis (CF) is a hereditary disorder caused by mutations in the cystic fibrosis transmembrane conductance regulator gene (*CFTR*). However, both intrinsic and extrinsic variables directly and/or indirectly associated with *CFTR* likely influence the course of CF, particularly the immune/inflammatory phenotype of CF lung disease (1). Many pathological hallmarks of CF such as chronic airway infection, persistent inflammation (2), and defective mucociliary clearance, are consequences of deficient or defective CFTR protein in airway epithelial cells. Over time, these cells fail to eradicate pulmonary pathogens, contributing to a mucosal immunodeficiency syndrome (3–5). In previous studies, plasma from individuals with CF compromised biological signaling and dysregulated mRNA and miRNA interactions in peripheral blood mononuclear cells (PBMCs), suggesting an impaired response in the circulating immune cells of CF patients (6–10).

In support of this notion, innate immune cells, represented primarily by monocytes, macrophages and dendritic cells, which are initially recruited to combat bacterial pathogens, have been shown to be dysregulated in CF airways (10). Although the significance and molecular basis of this dysregulation remains unclear, mutations in *CFTR* can affect not only the innate immune function of airway epithelial cells, but also alter innate immunity that contributes to recurrent and progressive infection in CF (11) (12, 13). Our previous work showed that CF F508del/F508del (homozygous) murine macrophages have a defective response and reduced cytotoxic activity against the bacterial pathogen *Pseuodomonas aeruginosa* (*Pa*) (8) which is prevalent in CF patients (14, 15). While bacterial infections exploit these defects in macrophage function (16, 17), treatment with the CFTR modulator, ivacaftor, improves macrophage-mediated cytotoxicity (18). Further, long noncoding (lncRNAs) have key roles in regulating the innate immune response to *Pa* in CF, combining with other regulatory mechanisms to alter the expression of immune/inflammatory genes within monocytes and macrophages (19–22). However, the genes and regulatory pathways involved in this immune dysregulation in individuals with CF have not been well characterized.

As CF is an autosomal recessive disorder with two defective *CFTR* copies, heterozygous carriers are typically considered healthy. However, CF carriers have an increased risk for a broad range of conditions affecting multiple organ systems, including asthma and airway infections (23, 24). Several studies reported familial clusters of pulmonary infections with nontuberculous mycobacteria, suggesting genetic risk factors, including *CFTR* mutations (25, 26). The observations that both CF carriers (CF parents) and CF patients are at higher risk of CF-related conditions than people without *CFTR* mutations suggest that distinct *CFTR*-related mechanisms are at play in both heterozygous and homozygous individuals. For the first time, this study directly addresses potential mechanisms in heterozygous individuals that contribute to this susceptibility (27, 28).

To understand the distinct molecular features and pathways contributing to known immune phenotypes in individuals with CF and in carriers, we assembled a cohort of parent-child trios (CF proband, father, and mother) and 20 unrelated healthy controls (HCs) without *CFTR* mutations (**Table 1, Fig 1**). We identified *CFTR*-related immune suppression in each trio subgroup through transcriptomic profiling using three cellular models: 1) a PBMC model of the impact of the *CFTR* mutation in immune cells; 2) a plasma model, where the donor PBMCs act as reporters of the immune microenvironment and compromised immune/inflammatory conditions in individuals with CF; and 3) a THP-1 cellular model that replicates the effects of this immune microenvironment on monocyte and macrophage function. Utilizing a previously established cell composition deconvolution method (8), we observed gene suppression in innate immune cells from *CFTR*-mutated PBMCs and from healthy PBMCs incubated with CF plasma. We determined that the abundance of mononuclear phagocytes correlates with CF clinical characteristics. Plasma samples from individuals within the trios were used in *ex vivo* cultures of THP-1 monocytes and macrophages to further characterize the transcriptomic profiles unique to CF patients and the profiles shared by CF trios. Consistent with these findings, our gene set enrichment analyses indicated impaired responsiveness of CF PBMCs and monocytes to lipopolysaccharide (LPS), an integral component of the *Pa* cell envelope. This approach of blood-based profiling and association with CF disease state provides important clues in understanding the vulnerability of carriers of *CFTR* mutations.

**Figure 1.**
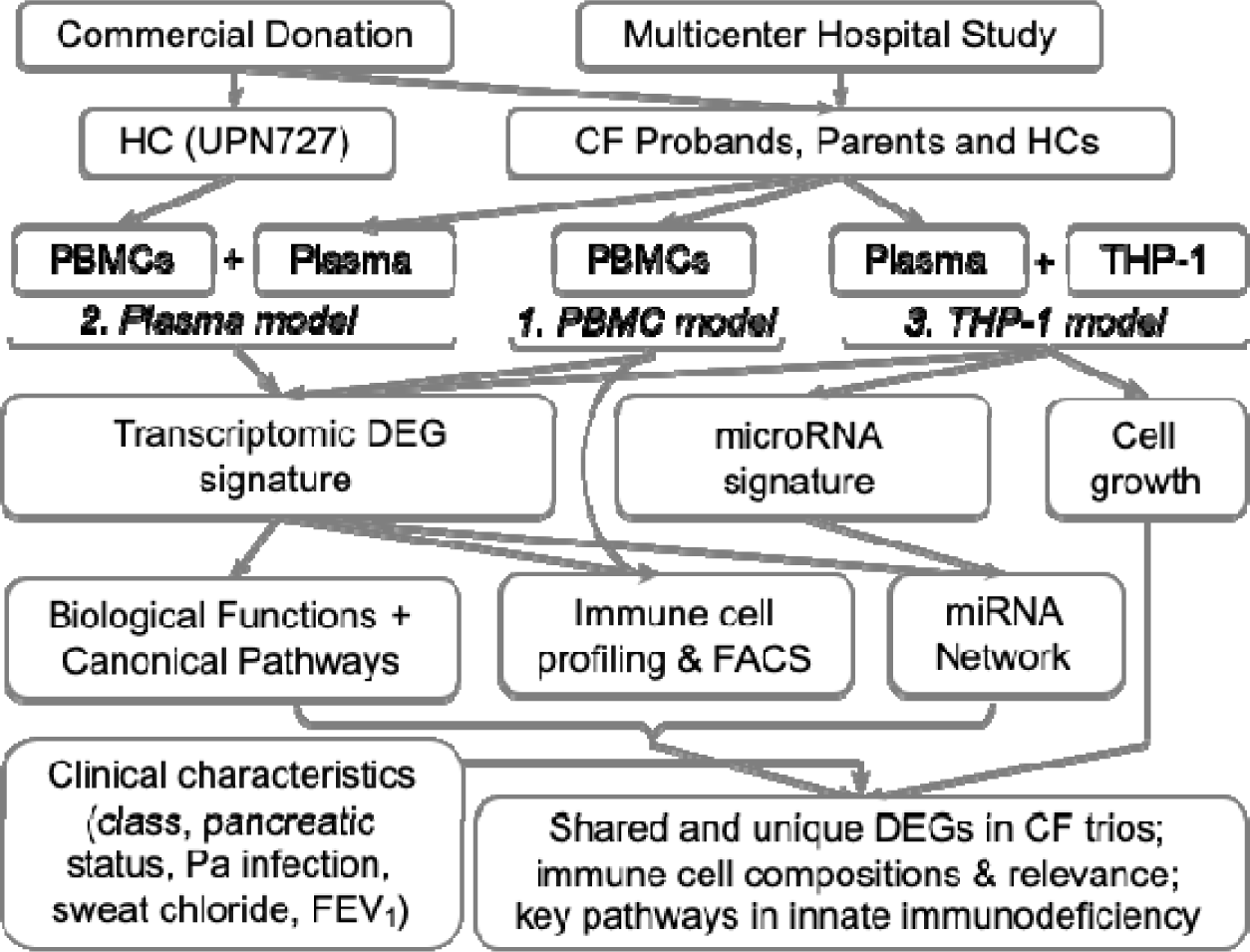
Schematic of main study procedures. See Table 1 for quantitative information about subjects in each model.

**Table 1.**
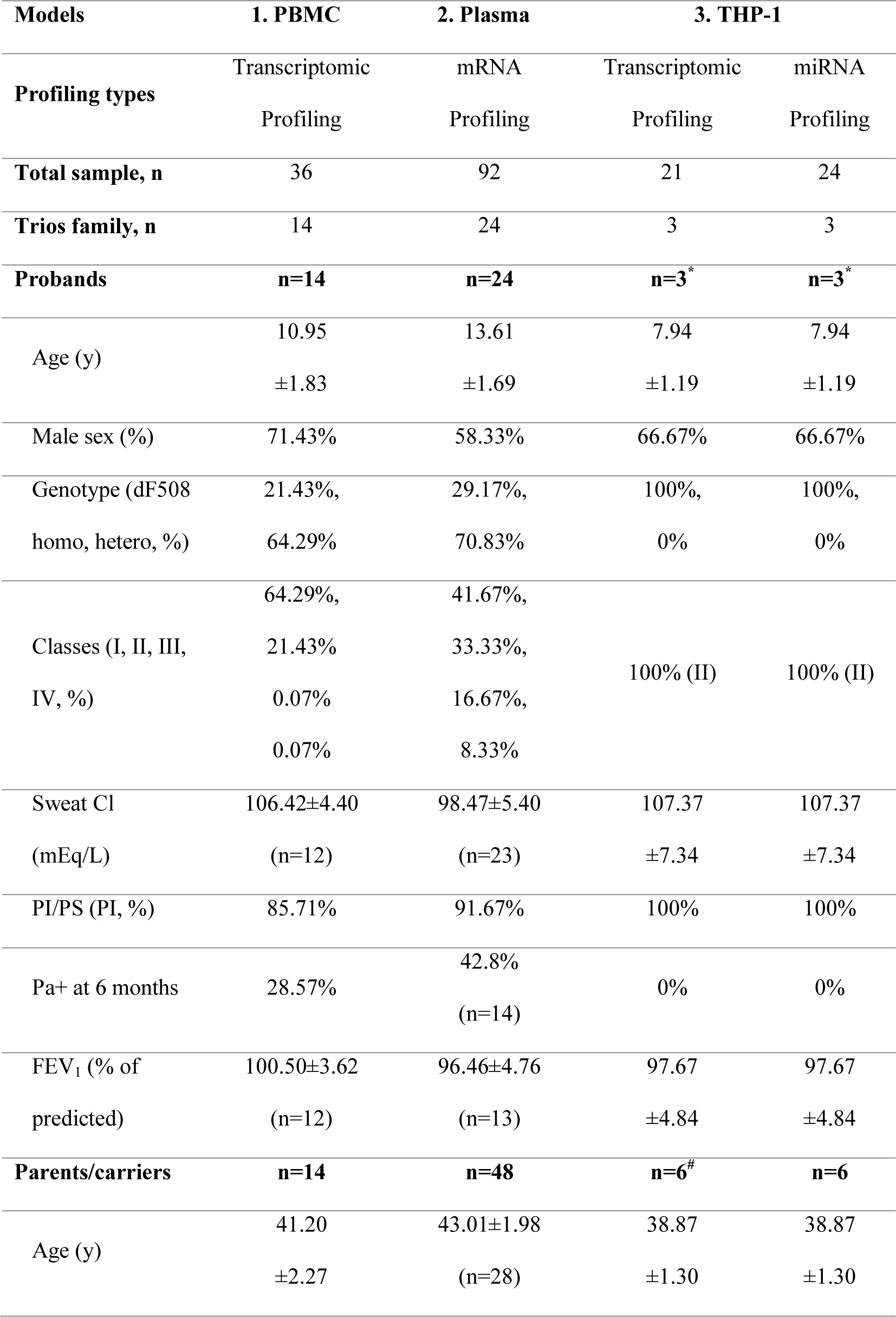
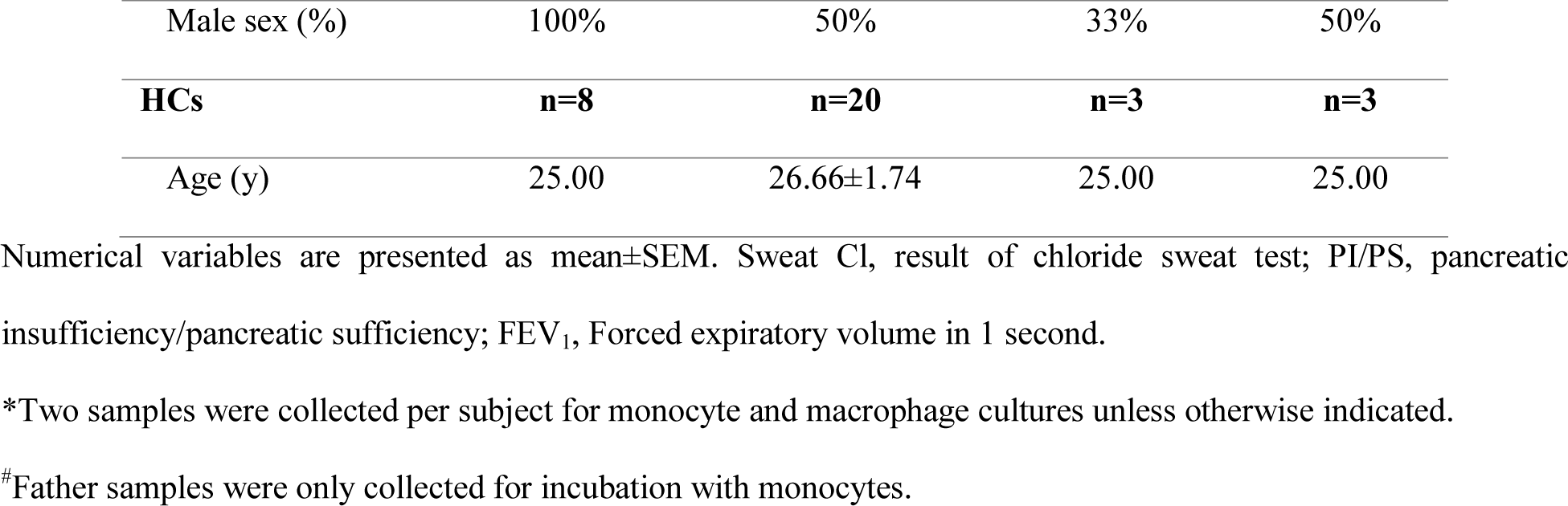
Demographic information of study subjects used for molecular profiling

## Results

### Profiling of PBMCs reveals significant downregulation of immune/inflammatory markers

To identify blood-based gene predictors that are clinically useful, we collected PBMCs from CF probands (n=14), their mothers and fathers (n=14), and HCs (n=8) and measured transcriptional expression in these cells with a whole-transcriptome array (Figure 1; Table 1, PBMC Model; Methods). Differential expression analyses revealed that a substantial number of transcripts (2267 out of 135,750) were differentially expressed between CF probands and HCs (Figure 2A, left). More than half of the differentially expressed genes/transcripts (DEGs) encode proteins (n=1422, including 213 from the “coding” category and 991 from the “multiple complex” category, where “multiple complex” is defined as a transcript reported in multiple locus types (Figure 2A, left; Table S1). Among these DEGs, >70% of coding RNAs (including multiple complex) and >80% of lncRNAs were downregulated in CF proband PBMCs (Figure 2A, right).

**Figure 2.**
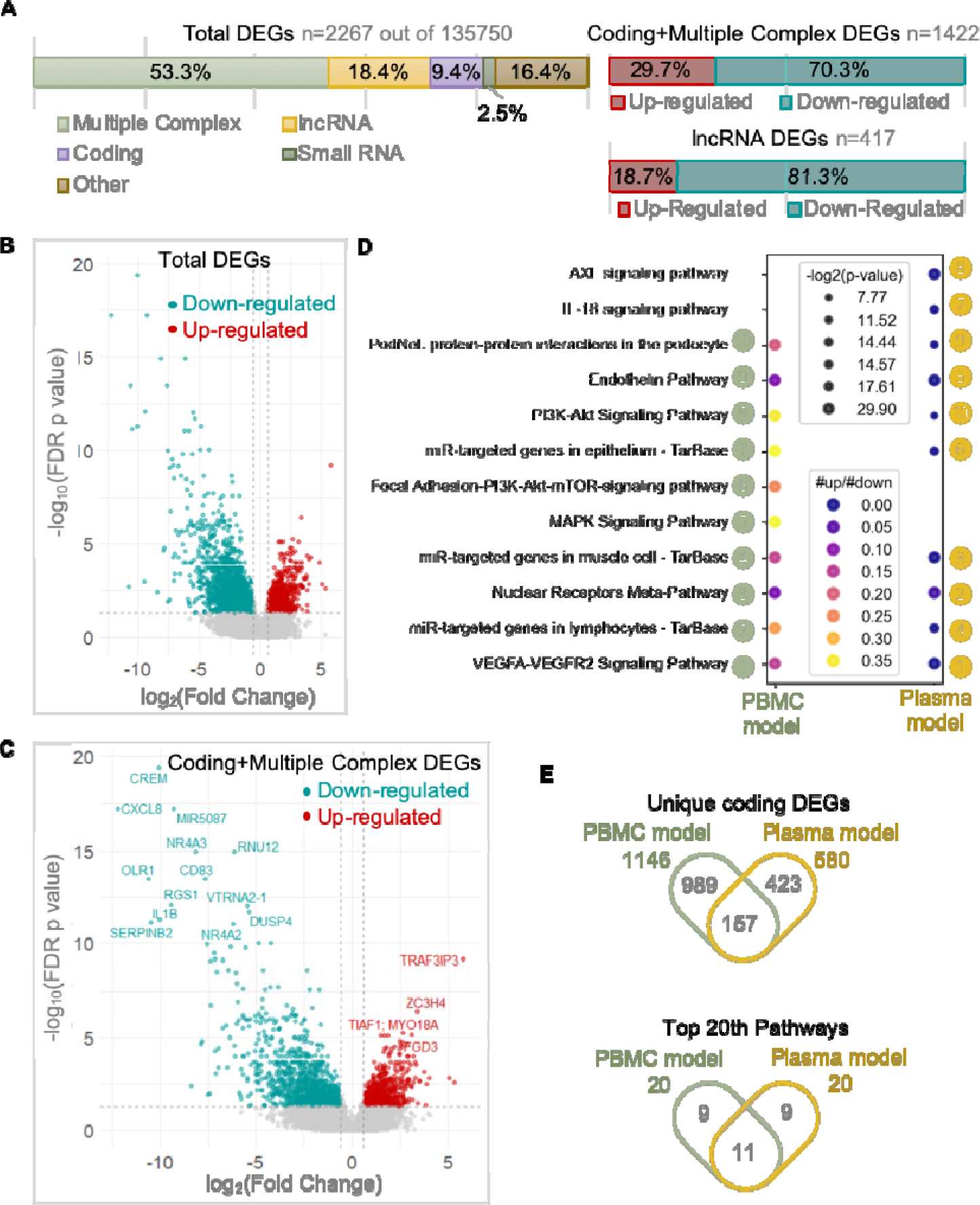
Immune-associated genes and pathways are significantly downregulated in CF PBMCs compared with HCs. **(A)** Transcripts differentially expressed between CF proband (n=14) and HCs (n=8) in the PBMC model, divided into categories according to locus type. We identified differentially expressed transcripts that displayed a more than two-fold change in expression level and a false discovery rate-adjusted p < 0.05. (**B-C**) Volcano plots of total differentially expressed transcripts and differentially expressed transcripts in the “coding” and “multiple complex” categories. **(D)** Bubble plot of the top 10 significant pathways in WikiPathways, ranked by the number of genes in the pathway, identified in the PBMC model (left side) and the plasma model (right side). For the plasma model: CF probands (n=24), HCs (n=20). **(E)** Venn diagrams showing the numbers and overlap of unique genes (upper) and top 20 pathways (lower) for CF probands versus HCs in the PBMC and plasma models.

We identified DEGs that showed large fold changes in expression in proband PBMCs relative to HC PBMCs (Figure 2B). Among these genes were key cytokines (e.g., *IL-1β*), chemokines (e.g., *CXCL8*), nuclear receptors (e.g., *NR4A3*), and modulators of immune signaling pathways (e.g., *CREM*) (Figure 2C). We identified fewer upregulated protein-coding genes than downregulated genes in the proband PBMCs, including proteins that interact with TNF receptor-associated factor (*TRAF3IP3*) and post-transcriptional regulators (zinc finger domain-containing protein *ZC3H4*) (Figure 2C). Most of the top-ranking pathways enriched in the DEG list were cytokine-related signaling pathways (29); each was downregulated in CF proband PBMCs compared with HC PBMCs (ratio of upregulated and downregulated gene numbers < 1) (Figure 2D). Thus, the observed imbalance between up- and downregulated transcript profiles in PBMCs of CF probands versus PBMCs of HCs provides further evidence of immune/inflammatory dysregulation in CF.

### CF probands and parents share similar transcriptomic features in PBMCs

To determine the transcriptomic signatures which were unique to and shared among the subgroups within our CF proband-parent trios, we performed transcriptomic profiling using both the PBMC and plasma models consisting of healthy donor PBMCs cultured with plasma from the CF probands or either parent (Figure 1; Table 1). The CF proband-associated transcriptomic profiles from the PBMC and plasma models shared common DEGs (Figure 2E; Figure S1; Table S2) and functional pathways (Figure 2E; Table S3), suggesting that compared with HCs, the CF probands exhibit extensive alterations leading to abnormal immune cell composition (Figures 4 and 5), as well as cytokine and chemokine profiles (Figure 2D).

Principal component analysis (PCA) of transcriptomic profiles from the plasma model (n=92 in 4 subgroups; Table 1) revealed overlap among the CF proband and parent subgroups within the CF trios, and which clustered separately from the HCs (Figure 3A; Figure S2A, left). In contrast, PCA of transcriptomic profiles from the PBMC model (n=36 in 3 groups) indicated no separation between the HC and CF trios subgroup clusters, although the HC cluster was not as broadly distributed as the proband and mother clusters (Figure 3A; Figure S2A, right). There was substantial overlap in the DEGs from CF probands versus HCs and from CF parents versus HCs in both the PBMC model (n=1737) and the plasma model (n=826) (Figure 3B). Interestingly, no significant DEGs were detected when comparing CF probands with parents in either model. In the PBMC model, we identified DEGs shared among CF probands and CF mothers (trios-shared genes; n=1737) and DEGs unique to CF probands (proband-unique genes; n=530), but no DEGs unique to CF mothers (Figure 3B; Tables S4-5).

**Figure 3.**
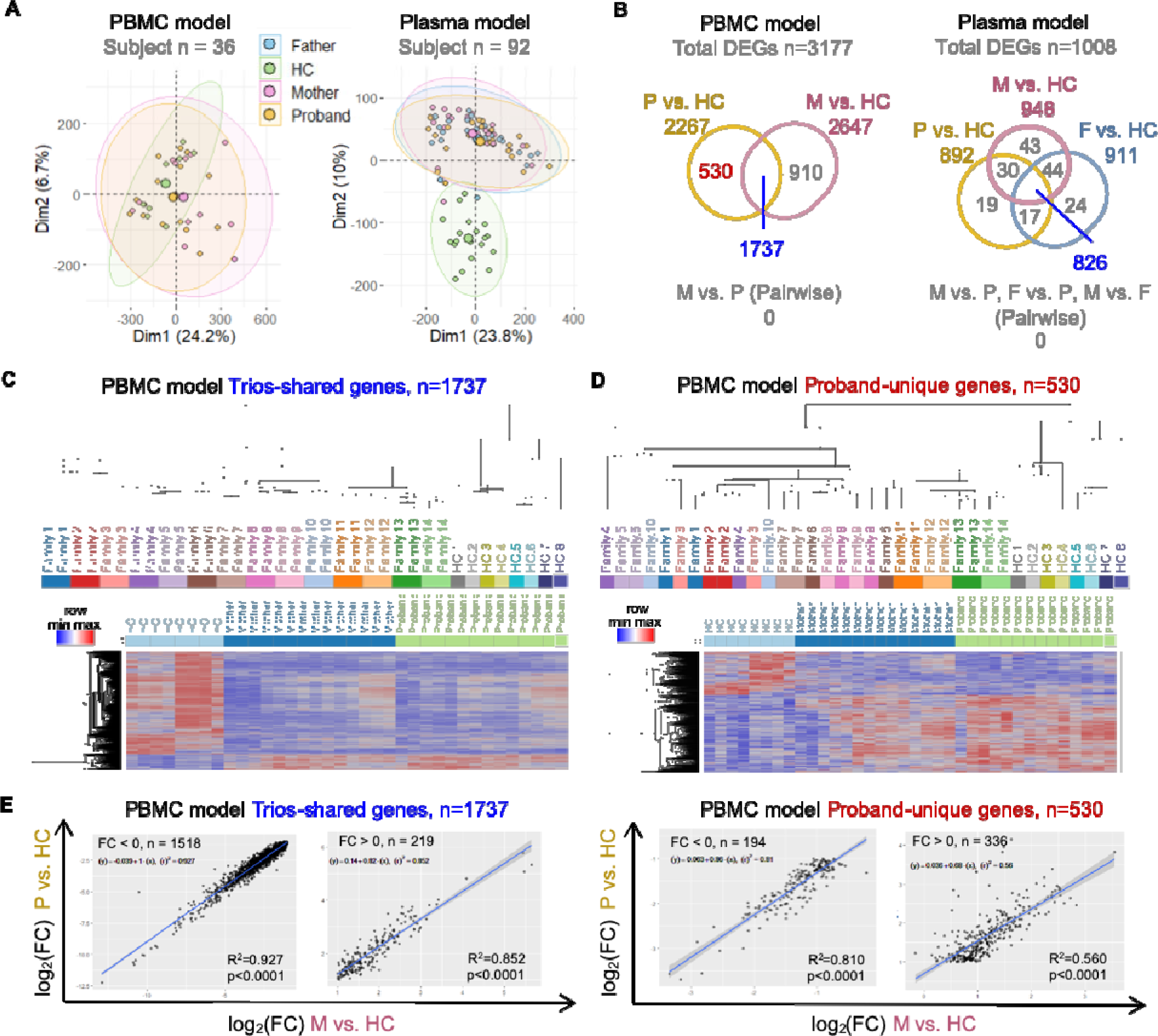
CF carriers and probands share highly similar transcriptomic profiles in PBMCs. **(A)** PCA of data from the PBMC (left, n=36) and plasma models (right, n=92). **(B)** Venn diagrams of the numbers and overlap of DEGs from the PBMC and plasma models; comparison are as indicated. Trios-shared genes and proband-unique genes are highlighted in blue and red, respectively. **(C-D)** Top, hierarchical clustering of study subjects; bottom, heatmap of the expression of trios-shared genes and proband-unique genes in the PBMC model. **(E)** Correlation scatter plots of the fold change (log_2_) of the two indicated comparisons of the expression of trios- shared genes (left) and proband-unique genes (right) from the PBMC model. The p value and R^2^ squared (square of the correlation coefficient) were produced by a Pearson correlation analysis. The linear regression line and its equation were generated from a simple linear regression analysis. P, proband; M, mother; F, father; FC, fold change.

We next performed hierarchical clustering predicting homogeneous groups among trio subjects. Notably, only the dendrogram generated using mother/proband-shared genes from the PBMC model (n=1737) organized subjects into groups in accordance with their family structures (Figure 3C, upper). This familial grouping was observed less consistently in the clustering dendrogram developed using proband-unique genes in the PBMC model (Figure 3D, upper) or trios-shared genes in the plasma model (Figure S2B). Overall, these results suggest that the mother/proband- shared genes in the PBMC model capture the relative homogeneity of gene expression across CF trios subjects.

The resulting heatmaps revealed near-identical expression patterns of trios-shared genes across the CF trios (mothers and probands vs. HCs; Figure 3C, lower). In contrast, we observed less similarity between mothers and probands in the proband-unique genes (Figure 3D, lower). We therefore asked whether the trios-shared genes displayed a consistent direction and magnitude of expression in the trios relative to HCs. The correlation between the expression levels of shared genes in CF probands and their mothers was significant (p<0.0001) and showed a strong, linear relationship (upregulated: R^2^=0.927, downregulated: R^2^=0.852) (Figure 3E, left). The correlation of the expression of proband-unique genes remained significant (p<0.0001) but showed a weaker linear relationship (upregulated: R^2^=0.81, downregulated: R^2^=0.56) (Figure 3E, right). Together, these transcriptomic profiling results indicate that parents and CF probands share highly similar expression patterns in the PBMC and plasma models.

### CF probands and parents share unique immune cell compositions

We previously developed a cell composition deconvolution method to estimate immune cell-type composition from gene expression data (8). Using this method, we found that myeloid cell subsets were less abundant while lymphoid cell subpopulations were more abundant in THP-1 cells incubated with CF proband plasma versus HC plasma (8). Here, we employed a similar deconvolution method (Methods, Figure S3-4) to infer the immune cell compositions of CF probands and their parents in our plasma model with donor PBMCs. We estimated a significantly higher abundance of 4 out of 5 lymphoid cell subsets (total T cells, CD4^+^ T cells, B cells, and natural killer cells) and a significantly lower abundance of all 5 myeloid cell subsets (monocytes, macrophages, activated monocytes, activated macrophages, and dendritic cells [DCs]) in cells incubated with CF proband plasma than in cells incubated with HC plasma (Figure 4A; Figure S5A). These differences were also observed when comparing results obtained with the plasma of CF parents compared to HC plasma.

**Figure 4.**
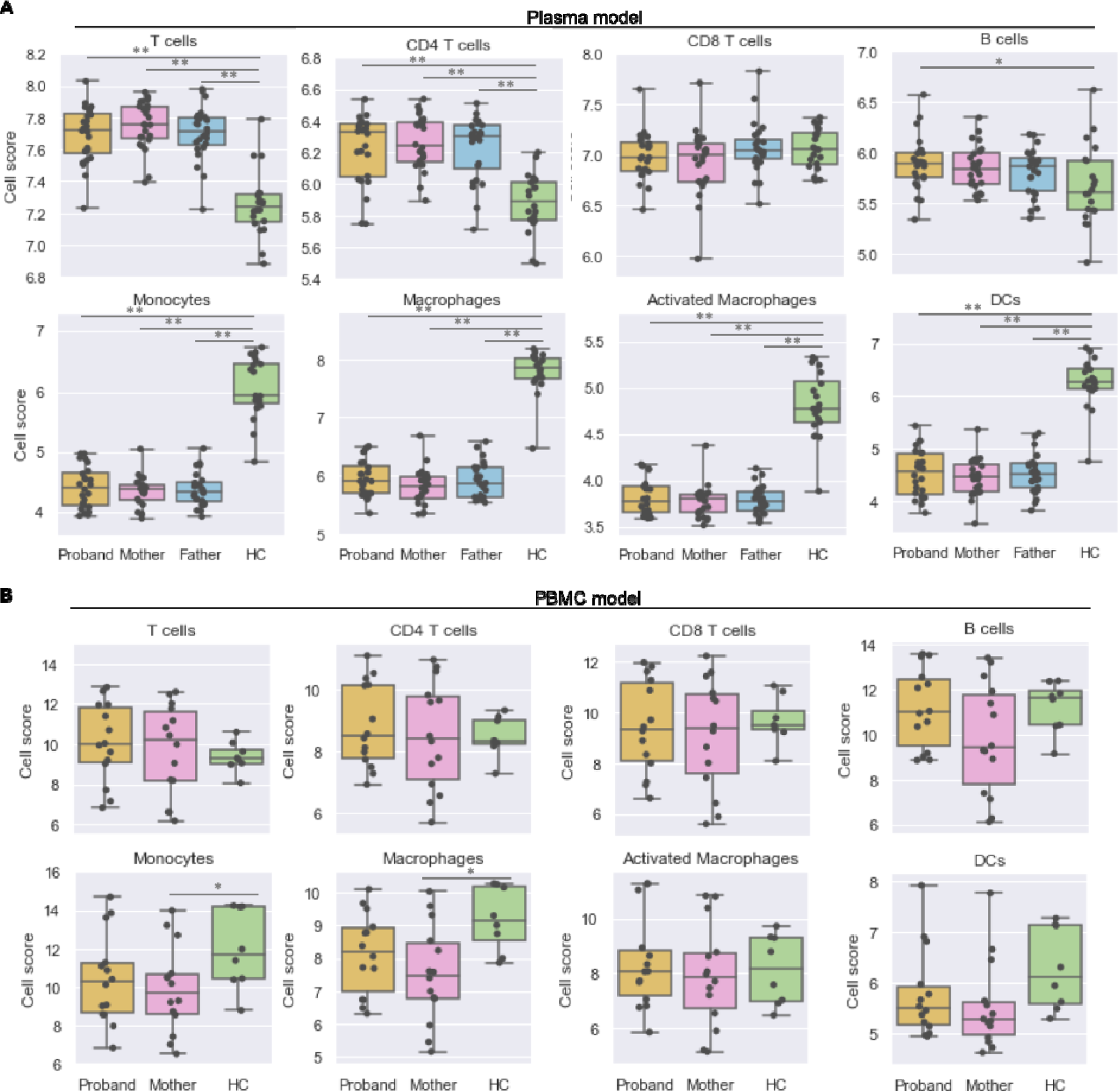
Immune-cell composition differs between CF parent-child trios and HCs. Dot plots and box plots of the cell composition scores of eight cell subsets in the **(A**) plasma model (probands, n=24; parents/carriers, n=48; HCs, n=20) and **(B)** PBMC model (probands, n=14; parents/carriers, n=14; HCs, n=8). Estimates of cell numbers in each cell subset were calculated for CF trios and HCs. The equality of variances was tested and confirmed by F-test and the normal distribution in each sample group was confirmed via Shapiro-Wilk normality test; the means of all comparison pairs were compared by unpaired independent t-test.*p <0.05, **p < 0.01.

The immune cells in the plasma model were harvested from a single donor (identical genomic background), whereas the immune cells in the PBMC model were harvested from the CF subjects and HCs (yielding greater genomic heterogeneity; Table 1). We hypothesized that mutations in *CFTR* might lead to a phenotype that disrupts immune-cell composition in CF PBMCs. Indeed, higher and lower cell numbers, respectively, were estimated in the lymphoid and myeloid cell subsets of PBMCs from CF probands versus HCs, but these differences were not significant (Figure 4B; Figure S5B). Monocytes and macrophages were significantly less abundant in CF parents (p<0.05) than in HCs (Figure 4B), but no significant difference was seen in CF probands versus HCs. These findings were confirmed via flow cytometry: both total circulating monocytes (CD14^+^) and classical monocytes (CD14^+^CD16^−^) were less abundant in CF probands than in HCs (Figure S5D). These results suggest that immune cell composition differs between CF trios and HCs, potentially identifying involvement of the monocyte-macrophage lineage in CF-related immunodeficiency.

### Phagocytic cell abundance is associated with CF disease severity and progression

Mutations in *CFTR* are divided into 6 classes (I-VI) according to aspects of CFTR biogenesis, metabolism, and function (30). The clinical severity and progression of CF can be predicted by categorical attributes such as *CFTR* class, pancreatic sufficiency status, *Pa* infection status, and numerical measures such as sweat-chloride level and forced expiratory volume in one second (FEV_1_) (30–33). To associate our findings on immune-cell composition in CF with clinical parameters, we assigned CF probands into subgroups based on the categorical attributes listed above and related the cell-abundance scores to each subgroup. For both the PBMC and plasma models, significantly fewer monocytes and macrophages were seen in the severe CF subgroups (class I/II/III and pancreatic insufficient [PI]) than in the mild/moderate subgroups (class IV and pancreatic sufficient [PS]) or HCs (Figure 5A-D). *Pa* infection was negatively associated with monocyte abundance in the plasma model (Figure 5E) but not in the PBMC model (data not shown). Moreover, in the PBMC model the abundance of mononuclear-phagocyte subsets (macrophages, monocytes, and activated DCs) was negatively correlated (p<0.05) with sweat- chloride levels and FEV_1_ values (Figure 5F-G). Collectively, these analyses indicate that the immune dysregulation associated with CFTR mutations likely results from the loss of innate immune cells and that the deficiency of mononuclear phagocytic cells, in particular, is closely linked to *Pa* infection, clinical severity, and the progression of lung function impairment in CF.

**Figure 5.**
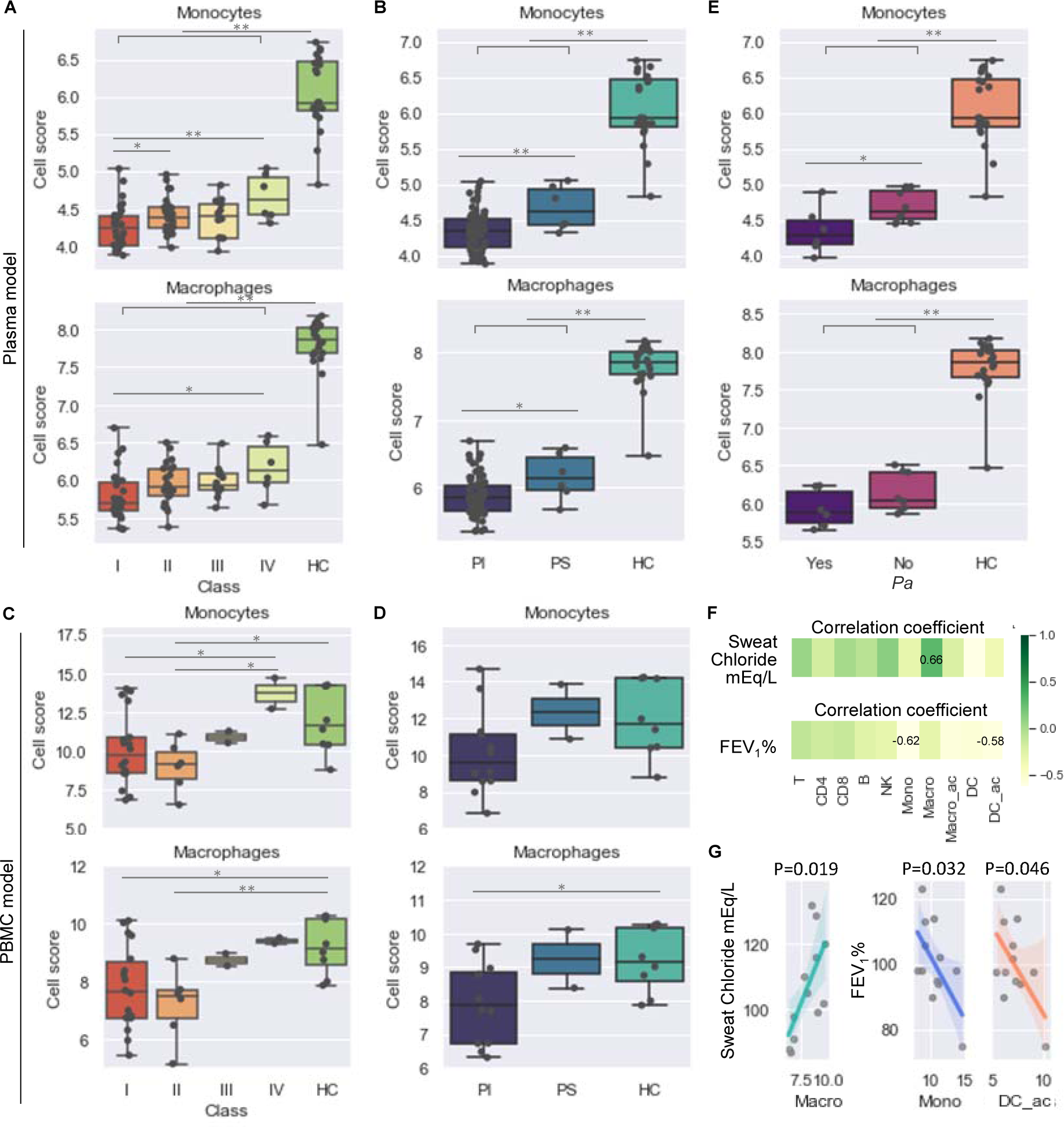
Compositions of monocytes and macrophages are correlated with CF disease severity and progression. **(A-E)** Dot plots and box plots of cell composition scores (Methods) of monocytes and macrophages in CF patient subjects grouped based on **(A, C)** their *CFTR* class, **(B, D)** pancreatic function, and **(E)** *Pa* infection status. Estimations of cell numbers in each cell subsets were compared between subgroups of CF patients. **(F-G)** Correlation analysis of cell abundance with sweat chloride and percent predicted FEV_1_. Plasma model, **(A, B, E)**; PBMC model **(C-D, F-G)**. The equality of variances was tested and confirmed by F-test; the normal distribution in each sample group was tested and confirmed by Shapiro-Wilk normality test; the means of all comparison pairs were compared by unpaired independent t-test (equal variances and normal distribution assumed). The p value and R (correlation coefficient) were produced by a Pearson correlation analysis (normal distribution assumed). *p <0.05, **p < 0.01. Mono, monocytes; Macro, macrophages; Macro_ac, activated macrophages; DC_ac, activated dendritic cells.

### CF trios plasma triggers a robust response in THP-1 monocytes but not macrophages

The monocyte cell line THP-1 has been used extensively to study monocyte and macrophage functions, mechanisms, and signaling pathways (34). Given the identification of monocytes and macrophages in the analyses reported above, we turned our attention to a model in which we cultured CF proband plasma with THP-1 cells in the presence or absence of phorbol 12-myristate 13-acetate (PMA) to differentiate these cells into macrophages (Figure 1; Table 1). Transcriptomic profiling identified >4,800 DEGs (out of 135,750 transcripts analyzed) in THP-1 monocytic cells cultured with plasma from CF probands or HCs, but only 199 DEGs from THP- 1 macrophage-differentiated cells cultured in the same manner (Figure 6A). DEGs from the THP-1 monocytes were consistently associated with a significant higher fold change than DEGs from the THP-1 macrophages (Figure 6B-C). In contrast to the results described above, in which most DEGs in CF proband cells were downregulated in PBMCs (Figure 2B-C), similar numbers of DEGs were both up- and downregulated in the THP-1 monocytes and macrophages (Figure 6B-C; Table S1-2). Pathway analysis of DEGs from the THP-1 model (both monocytes and macrophages) identified enrichment in immunoregulatory pathways such as transcription factors, cytokines, and receptors (Figure 6D; Table S3). Very few genes or pathway signatures were shared between the THP-1 model and the plasma model (Figure 6E) or the PBMC model (data not shown), however, consistent with the THP-1 model being a unique model specifically representing monocytes (34). In cell-growth assays of undifferentiated THP-1 cultures, plasma from CF probands with *Pa* infection significantly inhibited the proliferation of these monocytes (Figure 6F). Thus, plasma from CF probands appears to act as a modulator in an immunoregulatory capacity, inducing a broad and strong activation of THP-1 monocytes that is much more robust than that detected in THP-1 macrophages.

**Figure 6.**
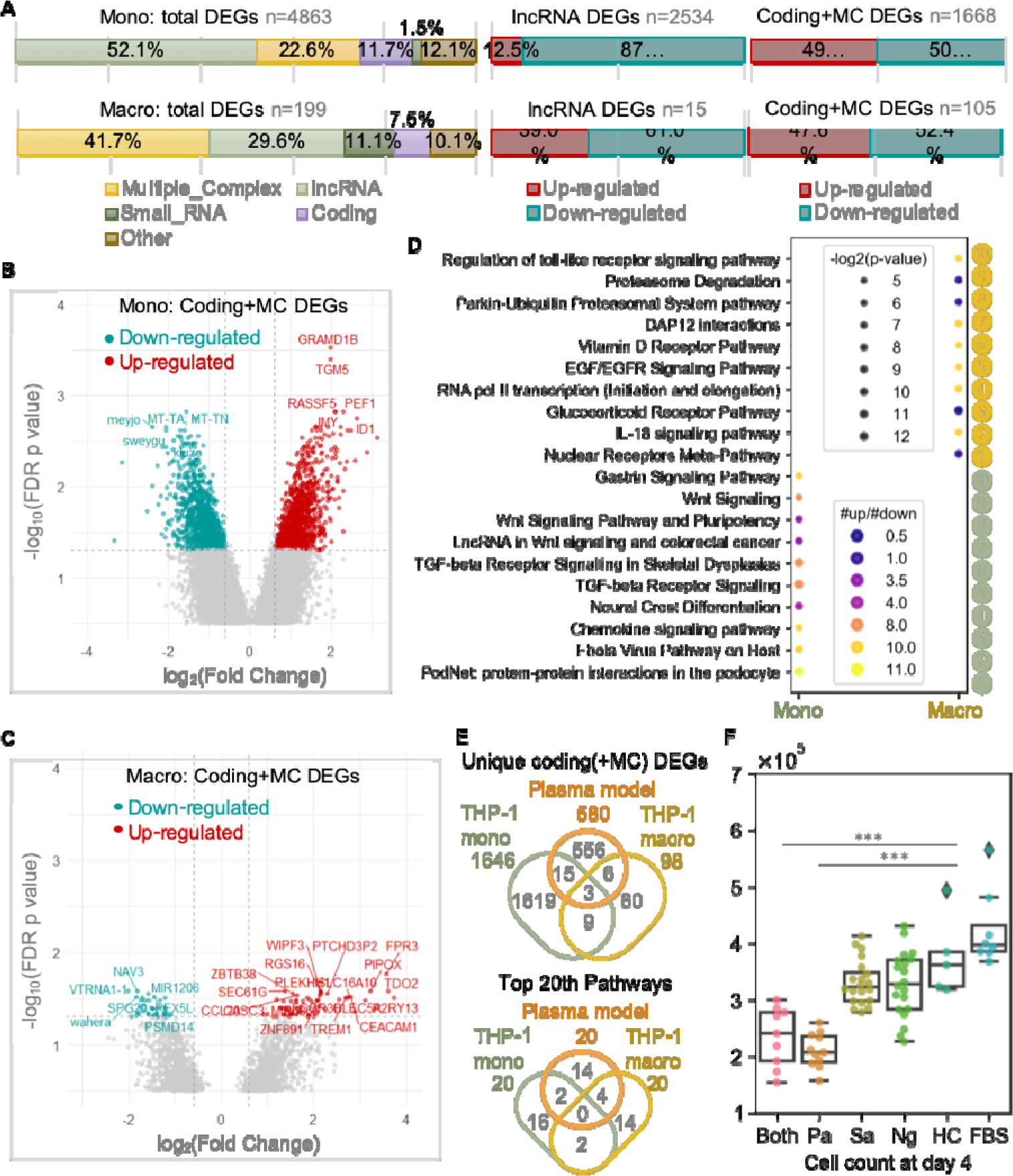
Plasma-cultured monocytes, but not macrophages, show dramatic changes in gene expression. **(A)** Breakdown of differentially expressed transcripts (fold change < -2 or > 2, false discovery rate p < 0.05, CF probands vs. HCs in the THP-1 monocyte and macrophage models) in main categories according to locus type. **(B-C)** Volcano plots of (**B**) total differentially expressed transcripts and the (**C**) differentially expressed transcripts in “coding” and “multiple complex” (MC) categories. **(D)** Bubble plot of top 10 significant pathways in WikiPathways ranked by number of regulated genes. **(E)** Venn diagrams of numbers and overlap of unique genes (upper) and top 20 pathways (lower) for CF probands versus HCs in the indicated models. **(F)** Bar plot of THP-1 cell numbers after 4 days of culture with CF plasma. Dunn’s multiple comparison for nonparametric post hoc testing was performed following the Kruskal-Wallis test to compare the differences between HC and other groups. ***p < 0.001. Pa, Pseudomonas; Sa, *Staphylococcus aureus*; Ng, negative; FBS, fetal bovine serum.

Next, we sought to determine whether the plasmas of CF probands and their parents evoke similar transcriptomic results in the THP-1 model. PCA of transcriptomic profiling data from both THP-1 monocyte and macrophage samples (n=21 in 7 groups) revealed that monocytes, but not macrophages, from CF trio subgroup clusters always overlapped, whereas the HC clusters remained distinct (Figure 7A). In the THP-1 monocytes and macrophages we identified trios- shared genes (n=1227 and 108, respectively) and proband-unique genes (n=2066 and 91, respectively) (Figure 7B, Table S4-5), as we did in the PBMC model (Figure 3B). As in the PBMC model, approximately one-fourth (1227 out of 4863) of the DEGs identified by comparing THP-1 monocytes incubated with CF proband plasma with those incubated with HC plasma were also differentially expressed in THP-1 monocytes incubated with CF parent plasma compared with those incubated with HC plasma (Figure 7B). Trios-shared genes and proband- unique genes in the THP-1 monocyte model were similarly identified from the total DEGs and used to identify homogeneous groups of subjects through hierarchical clustering (Figure 7C-D). Our analyses confirmed that trios-shared genes reflected familial structures and the expression patterns shared between THP-1 monocytes incubated with CF proband or parent plasma.

**Figure 7.**
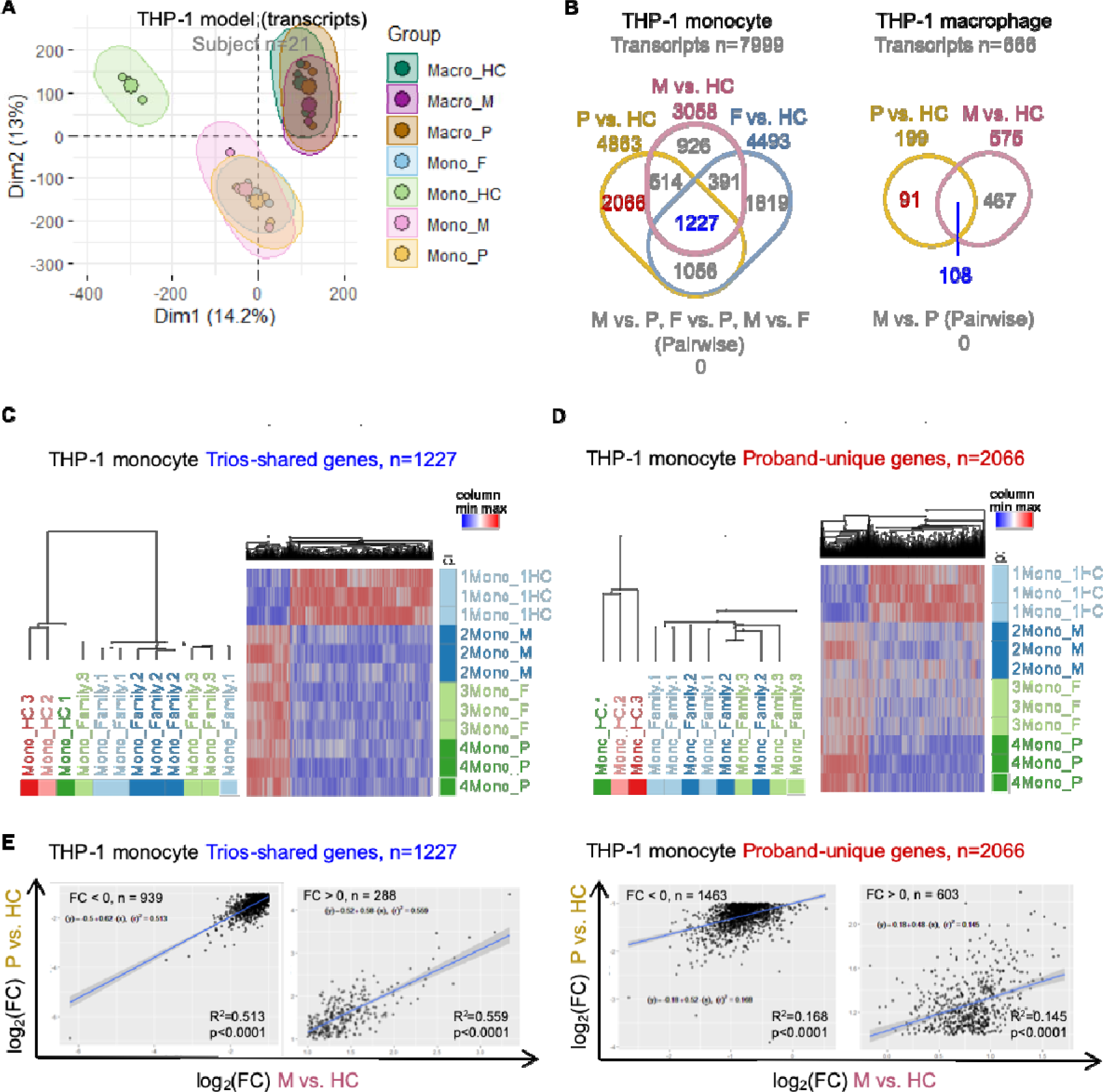
Transcriptomic profiles of CF carriers and probands are less correlated in the THP-1 monocyte model than the PBMC model. **(A)** PCA of study subjects in THP-1 monocyte and macrophage models based on similarities in transcriptomic profiling. **(B)** Venn diagrams of numbers and overlap of DEGs from THP-1 models. Trios-shared genes and proband-unique genes are highlighted in blue and red, respectively. **(C-D)** Hierarchical clustering of study subjects and heatmap of gene expression of **(C)** trios-shared genes and **(D)** proband-unique genes from the THP-1 monocyte model. **(E)** Correlation scatter plots of fold changes (log_2_) of the indicated comparisons of the expression of trios-shared genes (left) and proband-unique genes (right) in the THP-1 monocyte model. The p value and R^2^ squared (square of the correlation coefficient) were produced by a Pearson correlation analysis. The linear regression line and its equation were generated from a simple linear regression analysis. Mono, monocyte; Macro, macrophage; P, proband; M, mother; F, father; FC, fold change.

Moreover, the expression of trios-shared genes showed a significant (p<0.0001) and moderately linear relationship (upregulated: R^2^=0.513, downregulated: R^2^=0.559) in THP-1 monocytes incubated with CF proband or parent plasma, whereas proband-unique genes showed weaker correlations in expression between THP-1 monocytes incubated with CF proband or parent plasma (upregulated: R^2^=0.168, downregulated: R^2^=0.145) (Figure 7D; Figure S6). When we profiled miRNAs using an independent microarray (Methods), we observed similar miRNA expression patterns across CF trio groups (Figure S7A) and miRNA profiling revealed no significant differences in the expression of genes or miRNAs between THP-1 monocytes incubated with CF proband or parent plasma (Figure S7B). Therefore, we conclude that plasma from CF parents induces an immune response very similar to that induced by plasma from CF probands.

### Endotoxin tolerance is involved in the immune response shared by CF trios subgroups

Given the similarities between results obtained with PBMCs and plasma from CF probands and parents, we sought to characterize the top-ranked upstream regulators and causal molecular networks in CF probands versus HCs (PBMC model) using Ingenuity Pathway Analysis (IPA) (Methods). Interestingly, we identified “lipopolysaccharide (LPS)” as the top-ranked upstream regulatory molecule and “LPS-associated signaling” as the top-ranked causal network inhibited in PBMCs with a *CFTR* mutation (Figure 8A; Table S6). Of note, LPS is an integral component of the *Pa* cell envelope (REF). To validate this finding using large collections of published studies, we identified two core sets of protein-coding genes in the DEGs from the PBMC and plasma models (n=140) and the THP-1 monocyte model (n=365) (Figure 8B; Figure S8A). Three input gene sets (Table S7) that included genes regulated in the same direction (up- or downregulated) were submitted for gene set enrichment analysis (GSEA) in the Molecular Signatures Database (MSigDB) (35–38). Searches on input gene sets #1 and #2 returned significant matches that included gene sets previously defined from PBMCs, monocytes, or macrophages (Figure 8C; Figure S8B). Out of the top 10 gene sets that matched input gene set #1, 5 were associated with LPS or TLR4-interacting protein triggering receptor expressed on myeloid cells-1 (TREM1; Figure 8C). Similarly, input gene set #2 matched with LPS-stimulated gene sets (Figure S8B).

**Figure 8.**
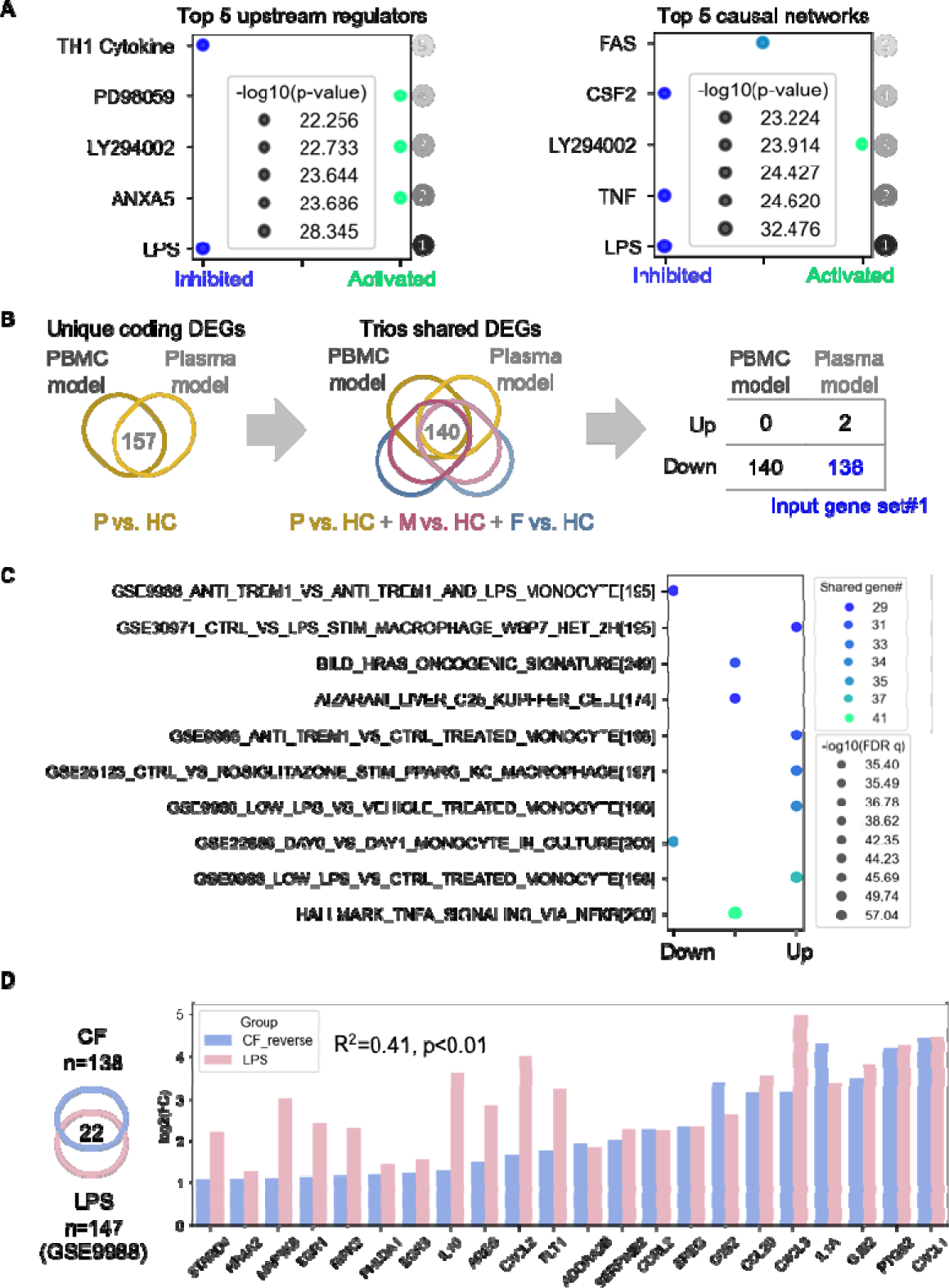
Integrated pathway enrichment analysis suggests an “LPS tolerant” state in CF trios. **(A)** Bubble plot of the top 5 significant upstream regulators and causal networks, ranked by p-value, from IPA using genetic profiles from the PBMC model (Methods). **(B)** Flow of identification and selection to identify input gene set #1 for gene set enrichment analysis. Firstly, the overlapping coding genes (n=157) from PBMC and plasma models (CF proband vs HC) were identified; then the overlapping genes (n=140) from these genes with DEGs from comparison of CF patients subjects versus HC were identified; the final input gene set (n=138) were identified as the genes regulated in same directions in both PBMC and plasma models. P, proband; M, mother; F, father. **(C)** Bubble plot of gene sets from gene set enrichment analysis matched with input gene set #1. Top 10 matched gene sets were ranked by q-value (false discovery rate). **(D)** Left, Venn diagram of the 22 overlapping genes from input gene set #1 and the annotated LPS-inducible gene set (GSE9988). Right, bar plot of fold change (log_2_) of the expression levels of genes in both input gene set #1 and the annotated LPS-inducible gene set (CF or LPS versus HC). Fold-change values from input gene set #1 were reversed from negative to positive for the convenience of visualization. The p value and R^2^ (square of the correlation coefficient) were produced by Pearson correlation analysis.

We next compared the fold changes in the expression of the LPS- and TREM1-induced genes shared in our profiling result with one published dataset from the search result (GEO GSE9988, CF proband versus HCs, THP-1 cells) (39). A significant but modest correlation was observed between genes identified in our experiment and the annotated LPS-induced genes (R=0.41, p<0.01) but not the TREM1-induced genes (R=0.001, p=0.88) (Figure 8D; Figure S8C). Together, these results suggest that monocytes from CF probands and carriers are less responsive to LPS, implying development of an LPS-tolerant state.

## Discussion

Recent work in our laboratory and others suggests a state of immune dysfunction in CF (4, 8), but it remains unclear whether this dysfunction arises from a primary intrinsic abnormality in immune cells themselves or is a byproduct of the infection microenvironment created by the *CFTR* defect. Abnormalities of *CFTR* function impair host defense, mucociliary clearance, and microbicidal activity in airways; dysfunctional immune cells contribute to the impaired response to infection(4). Consequently, most CF patients suffer intermittent infections with *Pseudomonas aeruginosa* (*Pa*) that progress to chronic infection(40). We previously demonstrated that CF patients exhibit changes in inflammation-related transcripts that correlate with disease status(6, 41), and we(8, 9) and others(18) identified monocyte and macrophage functions that were impaired in CF; these functions were not corrected by CFTR modulators.

In the present study, using 3 cellular models (Figure 1; Table 1) we capture the impact of the intrinsic deficiency (*CFTR* mutation) on circulating immune cells as well as the impact of the unhealthy immune microenvironment and compromised inflammatory milieu on circulating immune cells, particularly monocytes and macrophages, in CF probands and carriers. Our genomic and transcriptomic analyses identified DEGs that are associated with decreased activity in CF PBMCs. Our analyses also revealed changes in the abundance of mononuclear phagocytes (monocytes, macrophages, and DCs) associated with CF severity, *Pa* infection status, and disease progression. In addition, our THP-1 model demonstrated that monocytes—rather than monocyte-derived macrophages—respond dramatically to CF proband or carrier plasma. Gene-set enrichment analyses identified downregulation of LPS signaling in all cellular models, suggesting a loss of reactivity to LPS in CF PBMCs and monocytes.

Most existing CF profiling studies (42–46) have concentrated on CF airways and lungs, but CF immune dysfunction is likely extra-pulmonic, involving systemic alteration of immune function allowing for persistence of chronic infection (4, 11, 47). Consistent with this hypothesis, the immune-related transcriptomic profiles identified here (Figure 2A-2D) suggest that the innate immunodeficiency in CF is apparent outside of the chronically infected environment of the lung, a deficiency also seen in CF carriers.

In all 3 cellular models, we uncovered no significant differences between the transcriptomic profiles or those induced by the plasma of CF carrier and probands (Figure 3B, 7B), although we did identify groups of DEGs that were shared and co-expressed in CF patients and carriers (trios-shared genes) versus HCs (Figure 3C, 3E, 7C). These results are not surprising given prior evidence suggesting that a genetic load of 50% wild-type *CFTR* is not sufficient for maintaining health (24, 27). CF carriers may constitute a haploinsufficient population that is at a higher risk than the non-carrier population for developing respiratory infections and other diseases commonly associated with CF (23, 25–27).

PBMCs are a diverse mixture of highly specialized immune cell subsets that include myeloid and lymphoid cells (48). Under physiologic conditions in healthy subjects, CFTR protein is abundantly expressed in airway epithelial cells but expressed at lower levels in PBMCs (49). Nonetheless, CFTR is believed to carry out an irreplaceable function in myeloid cells (11, 18, 50). Notably, Sun and colleagues identified a set of genes in PBMCs that predict clinical responsiveness to ivacaftor therapy; using IPA, they mapped these genes to cellular processes that regulate innate immunity and inflammation (51). It remains unclear, however, whether these transcriptomic alterations were a direct effect of CFTR modulation in PBMCs or instead reflected systemic effects due to correction of the pathological environment generated as a result of the *CFTR* defect. Employing processes used to identify genes associated with ivacaftor responsiveness (51), we found that unlike HC PBMCs and HC plasma-cultured counterparts, the innate immune pathway was downregulated in *CFTR*-mutated PBMCs and in CF plasma-cultured healthy PBMCs and THP-1 monocytes (Figure 2D, 6D).

Monocytes typically account for 10–20% of total PBMCs found in blood (48), and these circulating monocytes are highly phagocytic (52, 53). Several powerful *in silico* approaches have been established to monitor changes in immune cell composition, using transcriptomic profiles to reveal distinct functionalities in cell subsets (54, 55). Our observations that monocytes and macrophages are significantly less abundant in CF trios subgroups than in HCs (Figure 4B) are consistent with the findings of our previous study (ref 8) as well as a recent report that the intrinsic molecular mechanisms controlling leukocyte recruitment and migration are severely impaired in CF monocytes (56). Taken together, these results support the notion that *CFTR* mutations lead to immune dysfunction and an immune deficiency. Notably, ivacaftor treatment does not change the abundance of PBMCs or the composition of immune subsets (monocytes, T cells, or B cells) (57). Further, these findings suggest that the cumulative abnormality in CF innate immunity is primarily caused by the fluid microenvironment resulting from the consequences of defective *CFTR* function. Since the intrinsic *CFTR* mutation disrupts monocyte recruitment and migration and cytokine levels and cell-to-cell interactions play major roles in this regulation (58), it follows that CFTR modulator therapy would not effectively reverse these consequences (59).

Given that individuals with CF commonly experience lung disease progression caused by chronic *Pa* colonization (14, 15), it is not surprising that *Pa* infection was associated with monocyte/macrophage abundance (Figure 5E), as observed in our previous study (ref 8). We also found evidence of negative associations between monocyte/macrophage abundance and CF disease severity (Figure 5A-D, 5F-G). Interestingly, these associations were not equally represented in the plasma and PBMC models: changes in monocyte/macrophage abundance were more strongly associated with *CFTR* mutation class (Figure 5A) and pancreatic status (Figure 5B) in the plasma model but more strongly associated with sweat chloride and FEV_1_ levels in the PBMC model (Figure 5F-G). These findings add to a growing body of evidence that the abundance of circulating monocytes can predict disease severity and progression as seen in chronic obstructive pulmonary disease (60) (61) (62) and idiopathic pulmonary fibrosis (63).

In our quest to identify the genes and pathways that underlie impaired innate immunity in CF, we uncovered a set of gene signatures (input gene set #1) that was shared by CF trios subgroups relative to HCs and downregulated in both PBMC and plasma models (Figure 8B). The gene signatures identified in the PBMC model (Figure 3C) better represent the features shared by CF trios than signatures identified from THP-1 cells (Figure 7C) because the correlation of the transcriptomic profiles between CF probands and carriers was much stronger in PBMCs (Figure 3E) than in THP-1 cells (Figure 7E). Further, GSEA revealed that these signature genes highly overlapped with LPS- and TREM1-induced genes, although these genes were regulated in opposite directions (Figure 8C-D). The significance of LPS or TREM1 as upstream regulators was supported by an independent IPA performed on data from the PBMC model (Figure 8A) and previous analyses of the plasma model (6, 8). As an activating receptor expressed on monocytes, TREM1 interacts and synergizes with the LPS/TLR4 receptor complex to trigger a respiratory burst, phagocytosis, and cytokine release in the innate immune system (39, 64, 65). However, CF monocytes are locked in an endotoxin-tolerant state (66) that is at least partly due to robust downregulation of TREM1 (67), and research has suggested a soluble endotoxin present in the bloodstream of individuals with CF may cause endotoxin tolerance in circulating monocytes (68). Consistent with these reports, we detected lower expression of LPS- and TREM1-induced genes (Figure 8C), suggesting that CF PBMCs are less responsive to LPS and have acquired an “LPS-tolerant state”. However, the presence of soluble endotoxin has not yet been independently verified, and the downregulation of TREM1 in CF patients has recently been challenged (69). How long does the tolerance to LPS last and is it reversible, given that CF patients can develop chronic *Pa* infection (70)? Better understanding of longitudinal exposure to LPS in CF patients may yield important mechanistic insights into the causes and consequences of LPS tolerance.

Although supportive of the concept of immune dysfunction and immunodeficiency, the present study includes a number of limitations. First, many differentially expressed lncRNAs were identified (Figure 2A, 6A) in both the PBMC and THP-1 models, but the clinical significance of these lncRNAs remains unclear, partly due to a lack of detailed information about their biological functions. Given that a significant number of lncRNAs has been implicated in LPS tolerance (71, 72), it remains unknown how their function enables long-lasting suppression of the LPS- and TREM1-induced pathways. Future research that includes longitudinal data, is needed to highlight lncRNAs with therapeutic potential for CF. Second, due to the small sample size in the PBMC model (Table 1) and the heterogeneity of human subjects, transcriptomic profiling failed to reveal transcripts that were significantly differentially expressed (Figure 3B, 7B) or were differentially spliced (data not shown) between a child with CF and either of their -parents. Third, no adjustment for multiple-hypothesis testing was performed in our correlation analysis between cell abundance and indicators of disease severity and progression (sweat chloride and FEV_1_ levels; Figure 5G). While a Bonferroni correction could have reduced the chance of a type I error, this approach was judged to be overly conservative because it hypothesizes that all variables (composition of immune cell subsets) are independent.

While homozygous mutations in *CFTR* cause CF, several studies suggest that heterozygous *CFTR* mutations have functional consequences (23). Our study has revealed similarity in the effects of *CFTR* mutation on innate immune cell populations in CF probands and carriers. CF carriers have an increased risk of developing airway obstruction (23, 73), neutrophil abnormalities, and ineffectual macrophage apoptosis (74). This study adds to the growing literature suggesting that the effects of heterozygous CFTR mutations on circulating monocytes could increase the prevalence of infection leading to chronic bronchitis and more severe lung disease (8, 75). The LPS-tolerant phenotype detected in this study may help explain the susceptibility present in carriers of homozygous and heterozygous *CFTR* mutations. Increased understanding of the role of novel lncRNAs and how they mediate signaling will provide clues for improved targeted therapies as research is beginning to confirm the role of lncRNAs as master regulators of gene expression.

## Methods

### Study subjects and data collection

A total of 164 CF-trios subjects (CF probands, mothers, and fathers) from 100 CF families and 20 unrelated healthy control (HC) subjects were recruited from the Children’s Hospital of Wisconsin (CHW 07/72, GC 390, CTSI 847, CHW 01-15), Ann & Robert H. Lurie Children’s Hospital of Chicago (2015–400), and National Jewish Health (NJH HS-3648) as this study (Figure 1) was approved by the Institutional Review Boards in each institution after scientific and ethical review. The Biomedical Research Alliance of New York (BRANY) is the Institutional Review Board providing oversight at National Jewish Health. HC subjects were free of known infection at the time of sample collection. Additional disease-free HC samples (n=11) were obtained commercially (Cellular Technology Limited, Cleveland, OH). Informed consent was obtained from subjects or their parents or legal guardians. As described in our prior studies (6, 8), CF proband subjects were diagnosed based on pilocarpine iontophoresis (CF Foundation guidelines) (76), symptoms, pancreatic status, *CFTR* mutation class, family history of CF, and information about the phenotypes of *CFTR* mutations (74, 77–79) (details in Supplementary Methods).

General demographic information, including age, sex, and genotype, was collected through standardized questionnaires. Pancreatic sufficiency status was defined based on levels of fecal pancreatic elastase, with a threshold of 200 μg/g for sufficiency (74). *Pa*-infection data was collected during standard screening for microbiological flora, in which the infection was reported as one positive microbiological growth from nasopharyngeal, sputum, or bronchoalveolar lavage specimens within 6 months of study enrollment. FEV_1_ data were collected during clinical lung function measurements performed at baseline according to ATS-ERS Task Force guidelines (80). Sweat chloride value was collected based on the sweat tests performed closest to the date of serum sample collection.

### Sample collection and cellular models

Human PBMCs or plasma samples from CF-trios subjects and age-matched, unrelated HCs were aseptically collected in acid citrate dextrose solution A or K^+^ EDTA anti-coagulant for the (Figure 1) the 3 cellular models for molecular profiling (Table 1). For the PBMC model, PBMCs (buffy coat) were collected from whole blood by Ficoll Paque (GE healthcare, Chicago, IL) density centrifugation. The PBMCs were then stored frozen in a cryoprotective medium containing 10% dimethyl sulfoxide (DMSO) and 90% fetal bovine serum (FBS). Cryopreserved PBMCs were thawed quickly before RNA isolation or live-cell recovery. The procedure used to develop the plasma model has been described in our prior studies (6, 8, 9). Briefly, healthy human PBMCs (UPN727; Cellular Technology Limited, Cleveland, OH) were co-cultured with the plasma collected from the enrolled subjects for 9 hours prior to sample collection for transcriptomic analyses. The THP-1 cell line was a gift from Dr. Peter H. Sporn (originally obtained from ATCC) and was maintained in RPMI 1640 medium supplemented with 10% FBS and 2 mmol/L L-glutamine. THP-1 monocytes were differentiated into macrophages by adding 200 nM PMA (Sigma-Aldrich, St. Louis, MO) to the media for 48 hours. The media was then removed and THP-1 monocytes or macrophages were co-cultured for 9 hours with plasma (without PMA) collected from the enrolled subjects.

Table 1 shows the numbers of CF trios and HC subjects used to develop the 3 cellular models. In addition to molecular profiling, cell samples from both the PBMC and THP-1 models were examined by cell-growth assays or fluorescence-activated cell sorting (Figure 1; Supplementary Methods). For cell-growth assays, THP-1 monocytes with no PMA treatment were cultured in media supplemented with FBS (n=3) and plasma from HC subjects (n=5) or CF probands that tested positive for *Pa* (n=13), positive for *Sa* (*Staphylococcus aureus*; n=22), positive for both (n=9), or negative for both (n=20).

### Molecular profiling and data processing

Total RNA was isolated using Trizol (Invitrogen, Waltham, MA) and the purity and concentration were verified using a NanoDrop ND-1000 instrument (Thermo Fisher Scientific, Waltham, MA). The integrity of the RNA was assessed by a 2100 Bioanalyzer gel image analysis system (Agilent, Santa Clara, CA). At least 300 ng mRNA per sample was submitted for library construction, in which each purified RNA sample was transcribed to double-stranded cDNA, followed by cRNA synthesis and biotin-labeling. The labeled samples were then hybridized onto three arrays (Table 1): GeneChip Human Genome U133 Plus 2.0 array (Thermo Fisher Scientific, >54,000 probes and >38,500 genes), Human Clariom D array (Thermo Fisher Scientific, >6,765,500 probes, >542,500 transcripts, and >134,700 genes), and GeneChip miRNA 3.0 array (Thermo Fisher Scientific, 1105 human miRNAs), as reported previously (6, 7). The profiling data from these arrays were normalized in the robust multi-array average (RMA) procedure, and then processed using Transcriptome Analysis Console (Thermo Fisher Scientific, version 4.0) following the manufacturer’s instructions.

### Immune cell profiling

Immune-cell profiling, or cell-composition analysis, was performed using a signature matrix optimized for human PBMC deconvolution, as reported in our prior study (8). Briefly, > 20 candidate marker genes for 10 cell subsets in PBMCs were selected from a previously described matrix based on their expression patterns across immune-cell subsets (82). The pairwise similarity statistic of all cell subsets (Figure S3-4) was computed between all pairs of the candidate marker genes within the normalized gene expression data from the PBMC model. Using the criteria (average Pearson correlation factor >0.50, p < 0.01), a number of selected marker genes were identified as our final marker genes (8). The raw cell-composition score was calculated as the sum of the simple averages of the marker genes’ log2 expression, which allows comparison of cell composition across subject groups and subgroups.

### Statistics

Bioinformatics and statistical analyses were performed and visualized using R version 3.6.1, Python version 3.7.9, Transcriptome Analysis Console (Thermo Fisher Scientific, version 4.0), Prism 7 (GraphPad, La Jolla, CA), IPA (Qiagen, Redwood City), and GSEA databases (Molecular Signatures Database, MSigDB version 7.2). Relative microarray gene-expression levels were compared between groups using an empirical Bayes method (ANOVA analysis followed by eBayes analysis) to share information across genes and generated an improved estimate for the variance. Coding genes and transcripts that displayed at least two-fold difference in gene expression between comparison groups (false discovery rate [FDR]< 0.05) were considered significantly differentially expressed and carried forward in the analysis. FDR adjustment was performed following Benjamini-Hochberg FDR-controlling procedure (83). Differentially expressed RNAs were illustrated as a volcano plot. Hierarchical clustering was performed to show the gene expression patterns and similarities among samples. PCA was performed to cluster subjects based on the differentially expressed transcripts. GSEA analysis was carried out by searching the established MSigDB gene-set collections (C1, C2, C7, C8) and utilizing the top 10 gene sets that matched with our input gene sets.

Associations between cell composition scores and clinical features were evaluated using Pearson correlation, R (correlation coefficient) and R^2^ (square of the correlation coefficient) (Figure 5). P-values in this analysis were not adjusted for multiple comparisons. Immune-profile scores were compared between groups and subgroups; independent t-tests were used when comparing CF trio samples with HCs, and paired t-tests were used when comparing between CF trio samples, assuming a normal distribution and equal variances. To compare the cell growth rates between groups, Dunn’s multiple comparison correction for nonparametric, post hoc testing was performed following the Kruskal-Wallis test. A p-value <0.05 was considered statistically significant.

### Study approval

This study was approved by the Institutional Review Boards (CHW 07/72, GC 390, CTSI 847, CHW 01-15 Children’s Hospital of Wisconsin, 2015-400 Ann & Robert H. Lurie Children’s Hospital of Chicago, NJH HS-3648 National Jewish Health), (Figure 1). The Biomedical Research Alliance of New York (BRANY) IRB is the current IRB providing oversight for this study at National Jewish Health. Written informed consent was received for each study participant. For all enrollees aged <18 years, a parent completed and signed the informed consent.

## Author contributions

XZ performed the experiments, built the data-analysis pipeline, analyzed the data, and wrote the draft of the manuscript. JD performed experiments and edited the manuscript. JK analyzed the data. The rest of the co-authors reviewed the manuscript and provided advice. HL devised the concept of the research incorporating the trio comparisons, supervised all aspects of the study, and reviewed and edited the manuscript.

## Supporting information

Supplementary Figures with Legends

## Data Availability

The datasets generated during and/or analyzed during the current study are available from the corresponding author on reasonable request.

## Acknowledgments

We thank all study subjects and their families who donated blood. We thank Dr. Peter H. Sporn (Northwestern University) for providing THP-1 cells. This work was supported by National Jewish Health Internal Grant support (to HL) and Crawford Foundation scholarship (to XZ).

## Supplemental Figure Legends

Figure S1. **Coding RNAs are significantly downregulated in donor PBMCs cultured with CF plasma. (A)** Differentially expressed mRNAs in CF probands versus HCs in the plasma model, divided into categories according to locus type. We identified differentially expressed transcripts that displayed more than a two-fold change in expression level and an FDR-adjusted p < 0.05. **(B)** Volcano plot of total differentially expressed mRNAs.

Figure S2. **PCA and hierarchical clustering of data from PBMC and plasma models. (A)** 3D PCA of clusters of study subjects in the PBMC (left, n=36) and plasma models (right, n=92). **(B)** Hierarchical clustering of study subjects using trios-shared genes from the plasma model.

Figure S3. **Pairwise similarity of predefined marker genes in 10 cell subsets from the PBMC model.** Pairwise similarity was computed based on gene expression across all subjects.

Figure S4. **Pairwise similarity of predefined marker genes in 10 cell subsets from the plasma model.** Pairwise similarity was computed based on transcriptomic expression across all subjects.

Figure S5. **Immune-cell composition associated with CF. (A-B)** Cell composition scores of natural killer (NK) cells and activated DCs in **(A)** the plasma model and **(B)** the PBMC model. The equality of variances was tested and confirmed by F-test; the normal distribution in each sample group was tested and confirmed by Shapiro-Wilk normality test; the means of all comparison pairs were compared by unpaired independent t-test, assuming equal variances and a normal distribution. **(C)** Cell-composition scores of monocytes and macrophages in (left) the plasma model and (right) the PBMC model. The means of each group were compared using paired t-tests (two subgroups) or ANOVA (three groups), followed by Tukey’s multiple comparison test. **(D)** Flow cytometry (left) of circulating total monocytes and classical monocytes in CF probands and HCs. Representative dot plots are shown on the right. Arrows represent gating strategy and process of selecting cell populations. Right, mean monocyte percentages of the 2 groups (HCs, n=3; CF, n=9). *p <0.05, **p < 0.01.

Figure S6. **Transcriptomic profiles of CF carriers and probands are moderately correlated in the THP-1 monocyte model.** Comparison of the fold changes (log_2_) of the expression levels of trios-shared genes in **(A)** probands and fathers versus HCs and **(B)** mothers and fathers versus HCs in the THP-1 monocyte model. The p-value and R^2^ (square of the correlation coefficient) were calculated using a Pearson correlation analysis. The line depicts the results of a linear regression analysis. P, proband; M, mother; F, father; FC, fold change.

Figure S7. **miRNA profiling of THP-1 monocytes incubated with CF proband or carrier plasma identifies limited signatures. (A) PCA of miRNA profiles from THP-1 cells incubated with study subject plasma. (A)** PCA of miRNA profiles from study subjects in the THP-1 model. **(B)** Venn diagrams showing the numbers and overlap of miRNA signatures from the THP-1 models. P, proband; M, mother; F, father.

Figure S8. **Pathway enrichment analysis supports an LPS-tolerant state in CF trios. (A)** Process of identification and selection to identify input gene sets #2 and #3 for GSEA. **(B)** Bubble plot of gene sets from GSEA matched with input gene set #2. The top 10 matched gene sets were ranked by q-value (FDR). P, proband; M, mother; F, father. **(C)** Left, Venn diagram of the 21 overlapping genes from input gene set #2 and the annotated TREM1-inducible gene set (GSE9988). Right, bar plot of fold changes (log_2_) in expression of the genes present in CF trios versus HCs, or TREM-1 versus HCs. Fold-change values from input gene set #2 were reversed from negative to positive for the convenience of visualization. The p-value and R^2^ (square of the correlation coefficient) were calculated using Pearson correlation analysis.

## Supplemental Tables

Table S1. All DEG lists identified by comparing CF probands and HCs.

• Table S1.1: DEG list from PBMC model (Human Clariom D Assay), n=2267

• Table S1.2: DEG list from plasma model (GeneChip Human Genome U133 Plus 2.0 Array), n=892

• Table S1.3: DEG list from THP-1 monocyte model (Human Clariom D Assay), n=4863

• Table S1.4: DEG list from THP-1 macrophage model (Human Clariom D Assay), n=199

• Table S1.5: DEG list from THP-1 model (GeneChip miRNA 4.0 Array), n=2 and n=118 for monocytes and macrophages, respectively

Table S2. Unique coding DEG lists identified by comparing CF probands and HCs.

• Table S2.1: Unique coding DEG list from PBMC model, n=1146

• Table S2.2: Unique coding DEG list from plasma model, n=580

• Table S2.3: Intersection of unique coding DEG lists from PBMC and plasma models, n=157 (related to Figure 2E)

• Table S2.4: Unique coding DEG list from THP-1 monocyte model, n=1646

• Table S2.5: Unique coding DEG list from THP-1 macrophage model, n=98

• Table S2.6: Intersection of unique coding DEG list from THP-1 monocyte, THP-1 macrophage and plasma models, n=3; intersection of THP-1 monocyte and THP-1 macrophage models, n=9; intersection of THP-1 monocyte and plasma models, n=15; THP-1 macrophage and plasma models, n=6 (related to Figure 6E)

Table S3. Significant pathways identified in WikiPathways by comparing CF probands and HCs.

• Table S3.1: Top 20 pathways from PBMC model ranked by total gene number

• Table S3.2: Top 20 pathways from plasma model ranked by total gene number

• Table S3.3: Intersection of the top 20 pathways of PBMC and plasma models n=11 (related to Figure 2E)

• Table S3.4: Top 20 pathways from THP-1 monocyte model ranked by total gene number

• Table S3.5: Top 20 pathways from THP-1 macrophage model ranked by total gene number

• Table S3.6: Intersection of the top 20 pathways from the THP-1 monocyte and macrophage models, n=2; THP-1 monocyte and plasma models, n=2; THP-1 macrophage and plasma models, n=4 (related to Figure 6E)

Table S4. Lists of trios-shared genes.

• Table S4.1: Trios-shared gene (DEG) list from PBMC model, n=1737

• Table S4.2: Trios-shared gene (DEG) list from plasma model, n=826

• Table S4.3: Trios-shared gene (DEG) list from THP-1 monocyte model, n=1227

Table S5: Lists of proband-unique genes.

• Table S5.1: Proband-unique gene (DEG) list from PBMC model, n=530

• Table S5.2: Proband-unique gene (DEG) list from THP-1 monocyte model, n=2066

Table S6. Significant upstream regulators and causal networks identified by comparing CF probands and HCs (PBMC model) using Ingenuity Pathway Analysis.

• Table S6.1: Top 5 upstream regulators ranked by p-value

• Table S6.2: Top 5 causal networks ranked by p-value

Table S7. Input and matched gene sets in the gene set enrichment analysis

• Table S7.1: Genes in input gene set #1 (n=138, downregulated)

• Table S7.2: Genes in input gene set #2 (n=163, upregulated)

• Table S7.3: Genes in input gene set #3 (n=202, downregulated)

• Table S7.4: Best-matched gene sets from MSigDB using input gene set #1

• Table S7.5: Best-matched gene sets from MSigDB using input gene set #2

