## Supplementary Figures with Legends for "*CFTR*-mediated monocyte-macrophage dysfunction revealed by cystic fibrosis proband- parent comparisons"

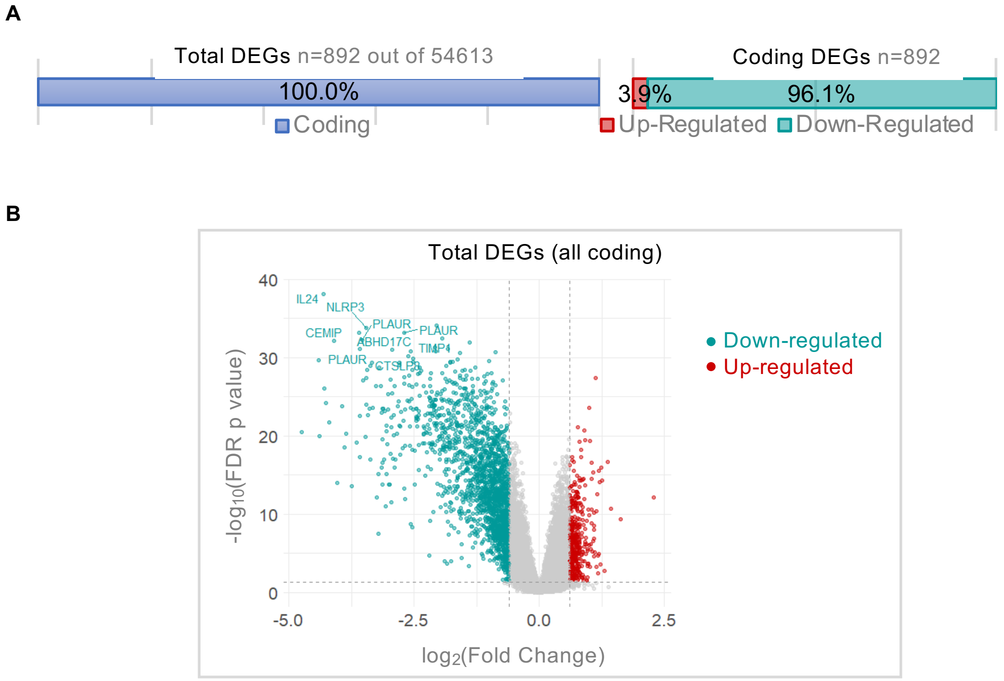


**Figure S1. Coding RNAs are significantly downregulated in donor PBMCs cultured with CF plasma. (A)** Breakdown of differentially expressed mRNAs (fold change < -2 or > 2, false discovery rate $p < 0.05$, CF proband vs. HCs in the plasma model) in main categories of locus type. **(B)** Volcano plot of total differentially expressed mRNAs.

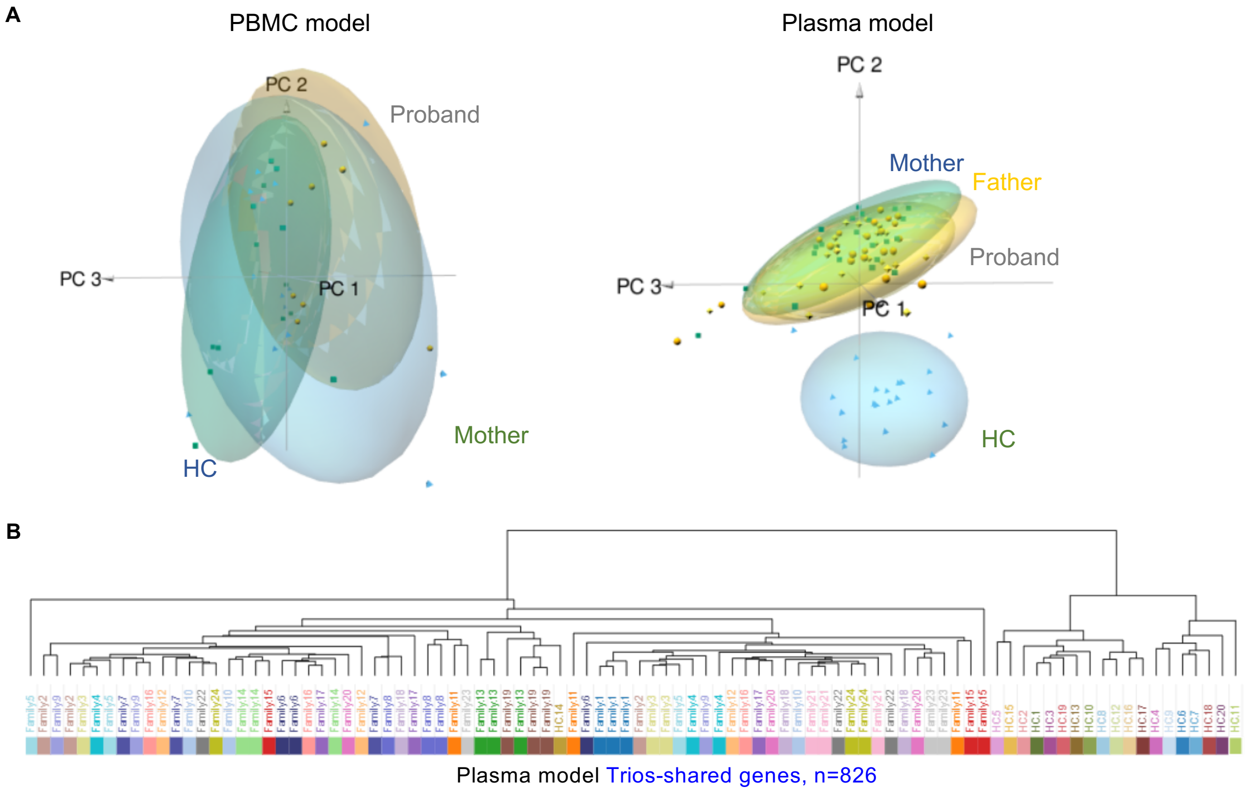


**Figure S2. PCA and hierarchical clustering of data from PBMC and plasma models. (A)** 3D PCA of clusters of study subjects in the PBMC (left, n=36) and plasma models (right, n=92). **(B)** Hierarchical clustering of study subjects using “trios-shared genes” from the plasma model.

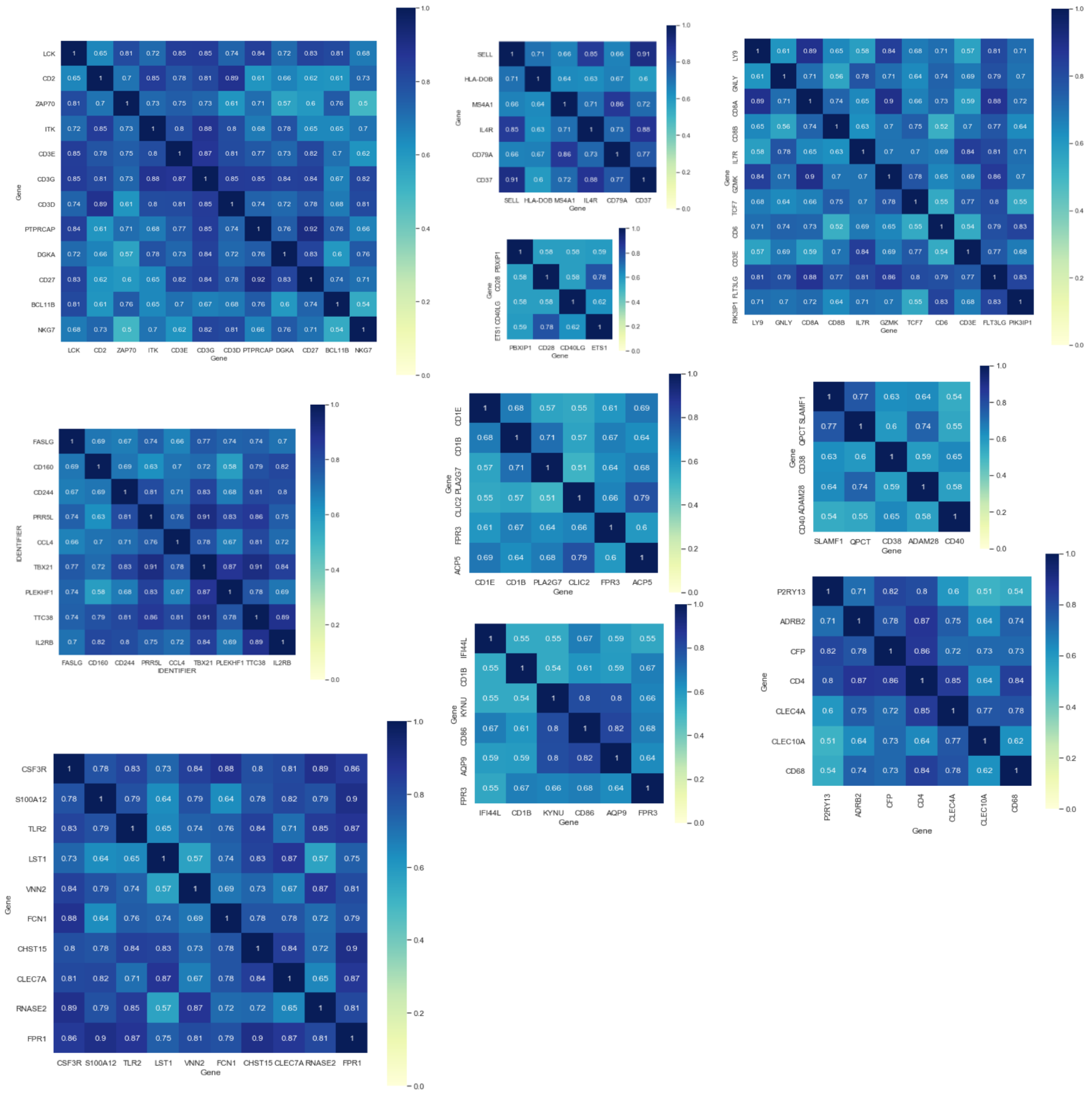

**Figure S3. Pairwise similarity of predefined marker genes in ten cell subsets from the PBMC model.** Pairwise similarity was computed based on transcriptomic expression across all subjects.

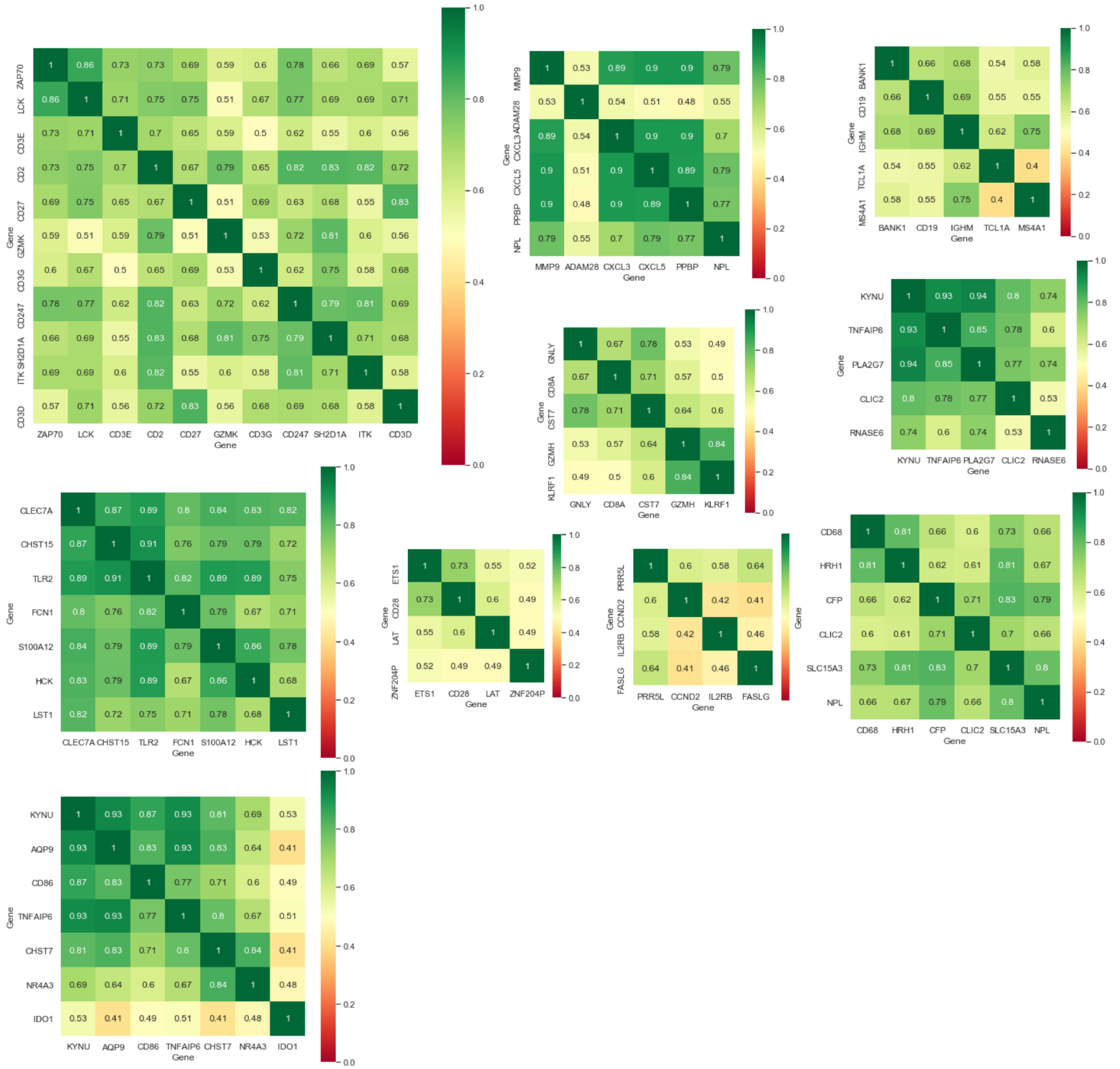


**Figure S4. Pairwise similarity of predefined marker genes in ten cell subsets from the plasma model.** Pairwise similarity was computed based on transcriptomic expression across all subjects.

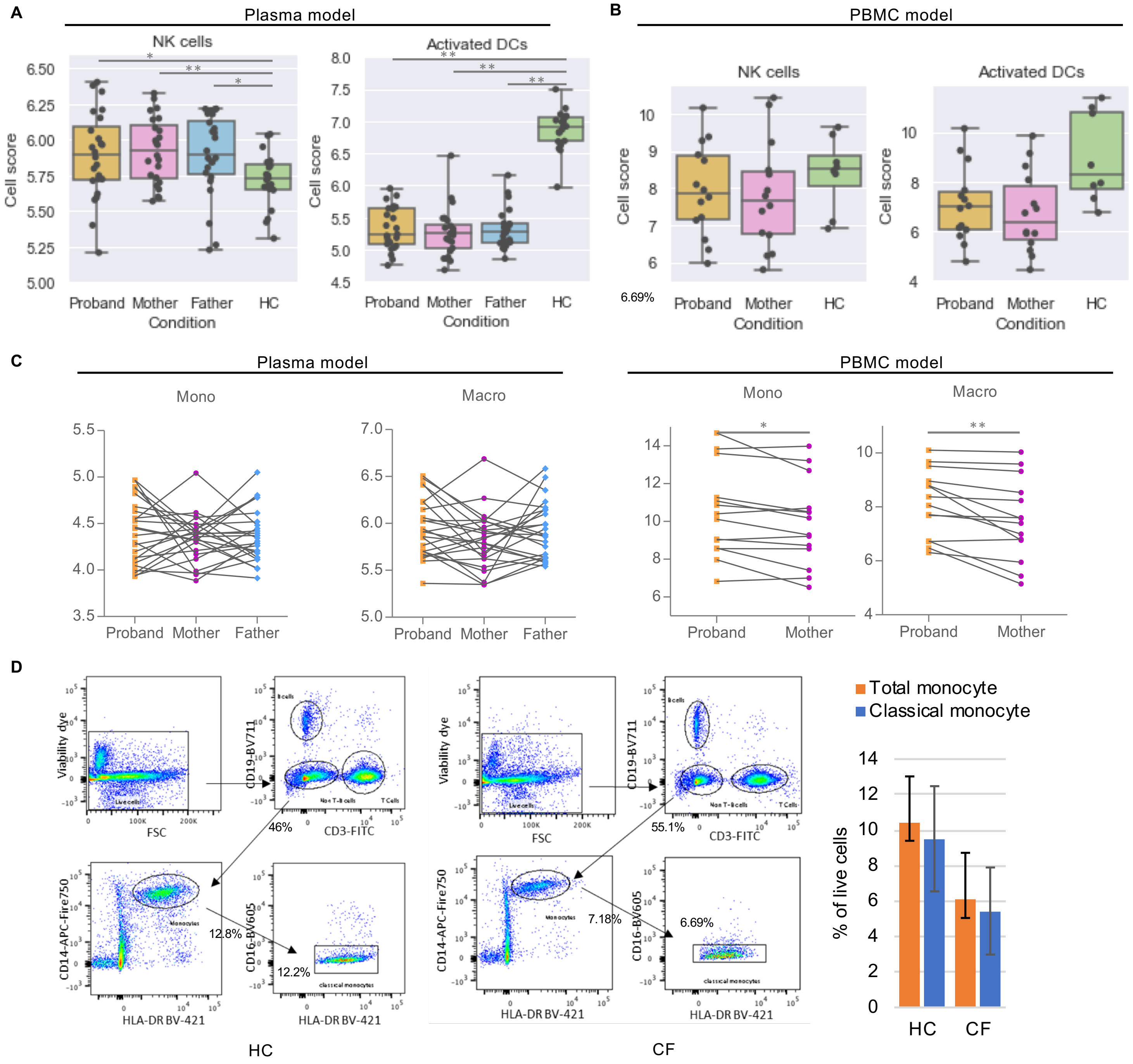


**Figure S5. Additional immune-cell composition analysis of CF parent-child trios and HCs. (A-B)** Dot plot and box plot of the cell composition scores of two additional cell subsets in **(A)** the plasma model and **(B)** the PBMC model. The equality of variances was tested and confirmed by F-test; the normal distribution in each sample group were tested and confirmed by Shapiro-Wilk normality test; the means of all comparison pair were compared by unpaired independent t-test (equal variances and normal distribution assumed). **(C)** Dot plot of the cell composition scores of monocytes and macrophages in (left) the plasma model and (right) the PBMC model. The means of all comparison pair were compared by paired t-test (two subgroups) or ANOVA (three groups) followed by Turkeys Multiple comparison. **(D)** Flow cytometry (Methods) of circulating total monocytes and classical monocytes in CF probands (left) and HCs (middle). Representative dot plots are shown. Arrows represent gating strategy and flow of selection of specific cell populations. Right, mean values of monocyte percentages of the two groups (HCs, n=3; CF, n=9). *p <0.05, **p < 0.01.

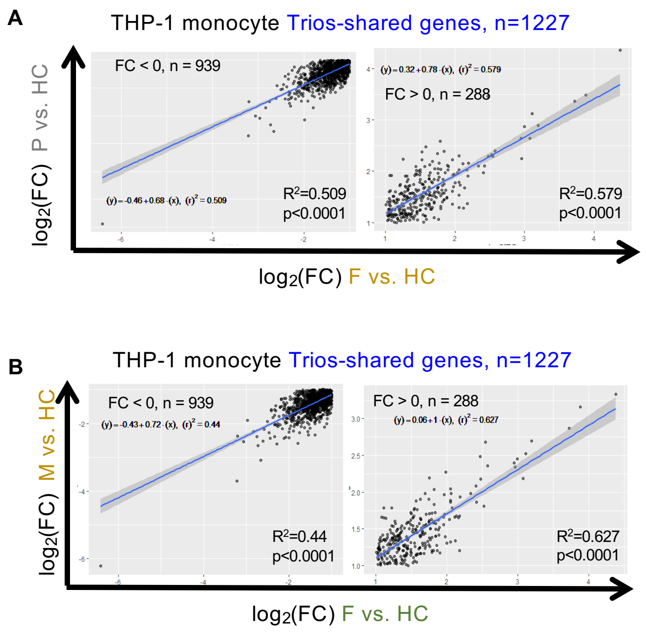


**Figure S6. Transcriptomic profiles of CF carriers and probands are moderately correlated in the THP-1 monocyte model.** Co-correlation scatter plots of the fold changes ($log_2$) of the expression levels of "trios-shared genes" in **(A)** proband and father versus HCs and **(B)** mother and father versus HCs in the THP-1 monocyte model. The p value and $R^2$ squared (square of the correlation coefficient) were produced by a Pearson correlation analysis. The linear regression line and its equation were generated from a simple linear regression analysis. P, proband; M, mother; F, father; FC, fold change.

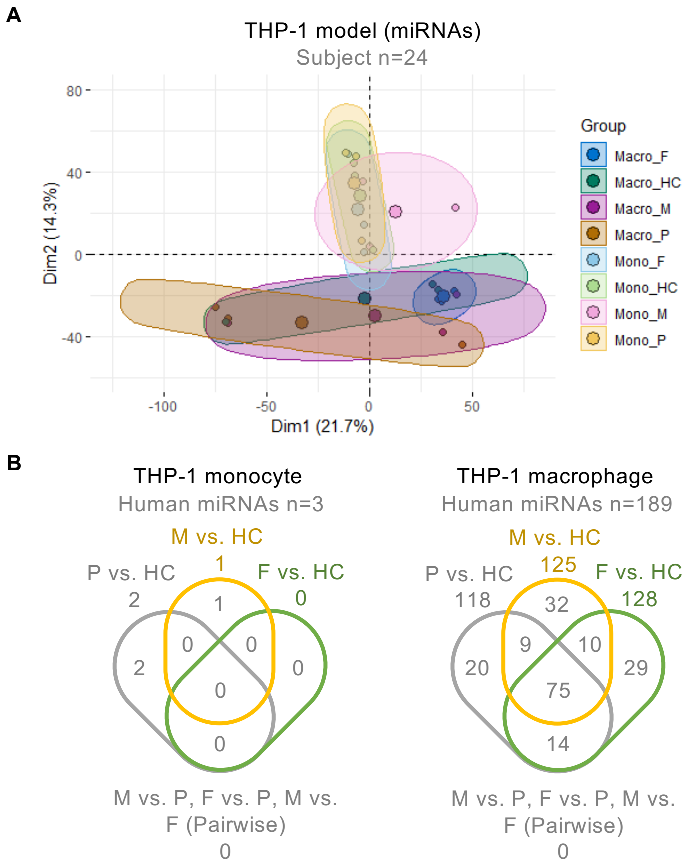


**Figure S7. miRNA profiling of CF carriers and probands identifies limited signatures in THP-1 monocytes. (A)** PCA of data from study subjects in THP-1 model based on their similarity in an independent miRNA profiling experiment (Methods). **(B)** Venn diagrams of numbers and overlap of miRNA signatures from the THP-1 models. P, proband; M, mother; F, father.

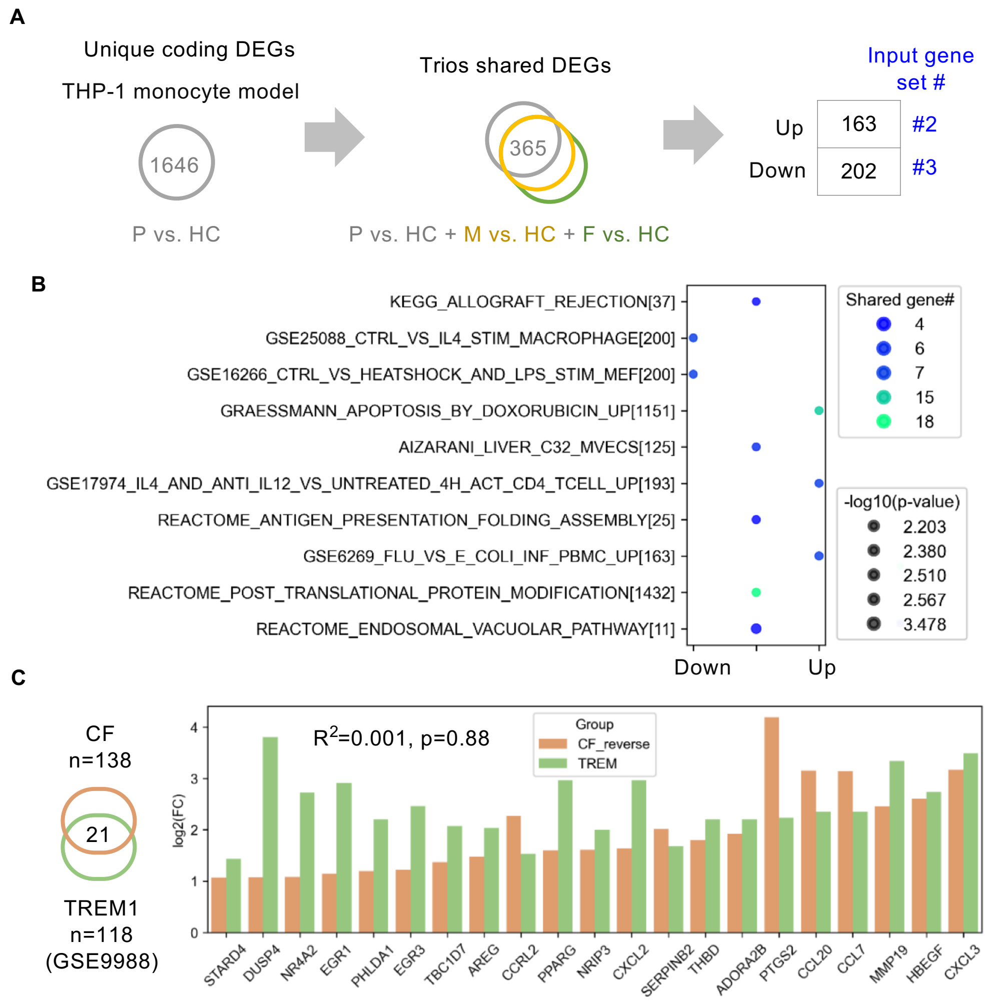


**Figure S8. Additional pathway enrichment analysis supports an "LPS tolerant" state in CF trios. (A)** Flow of identification and selection to identify input gene set#2 and set#3 for gene set enrichment analysis. **(B)** Bubble plot of gene sets from gene set enrichment analysis matched with input gene set#2. The top 10 matched gene sets were ranked by q-value (false discovery rate). P, proband; M, mother; F, father. **(C)** Left, Venn diagram of the 21 overlapping genes from input gene set#2 and the annotated TREM1-inducible gene set (GSE9988). Right, bar plot of fold change ($log_2$) of the genes presented in both gene sets (CF or TREM-1 versus HC). Fold-change values from input gene set# were reversed from negative to positive for the convenience of visualization. The p value and $R^2$ squared (square of the correlation coefficient) were produced by Pearson correlation analysis.
